# Statistical analysis for the development of a Deep Learning model for the classification of images with TDP-43 pathology

**DOI:** 10.1101/2024.02.12.24300689

**Authors:** Azucena Muñoz, Vasco Oliveira, Marta Vallejo

**Affiliations:** Heriot-Watt University, Edinburgh, UK; Francisco de Vitoria University, Madrid, Spain

**Keywords:** ALS, TDP-43, Deep Learning, Statistical Analysis

## Abstract

Diagnosing Amyotrophic Lateral Sclerosis (ALS) remains challenging due to its inherent heterogeneity. Cytoplasmic aggregation of TDP-43, observed in approximately 95% of ALS cases, has emerged as a key pathological hallmark. In this observational study, we investigated the feasibility of training deep learning models to classify TDP-43 proteinopathic samples versus healthy controls, with a particular focus on understanding how dataset limitations affect model performance.

The dataset comprised super-resolution immunofluorescence images in which cytoplasmic and nuclear TDP-43 deposits were quantified using red and pink pixel counts. We formulated three classification tasks: TDP-43 pathology (binary), TDP-43 pathology grades (multiclass), and ALS diagnosis (binary). Initial deep learning experiments yielded inconclusive results, prompting dataset curation and the removal of problematic samples.

Subsequent statistical analyses using t-tests, ANOVA, and hierarchical clustering revealed significant differences between healthy and pathological samples in terms of pixel distributions, total protein levels, and TDP-43 compartmentalisation. These findings suggest that classification based on TDP-43 proteinopathy provides a more reliable framework for deep learning compared to ALS diagnosis, underscoring the importance of data quality and task stratification in model performance.

## 1. Introduction

### 1.1. Amyotrophic Lateral Sclerosis

Amyotrophic Lateral Sclerosis (ALS) is recognised as a multisystem neurodegenerative disease with great clinical, genetic and neuropathological heterogeneity (1). The estimated incidence of ALS is 1.75 to 3 per 100,000 people per year, which rises to 4-8 in the highest-risk age group (45-75 years). Prevalence has significant geographic differences as it ranges between 10-12 per 100,000 people in Europe and 3.84-5.56 per 100,000 people in the United States. About 90-95% of cases are sporadic (sALS), and the remaining 5-10% are hereditary or familial (fALS). The estimated risk of developing ALS is 1:350 in men and 1:400 in women (2). Diagnosis primarily relies on phenotypic assessment as it is based on signs of upper motor neurons (UMN) or lower (LMN) motor neuron dysfunction in patients with progressive muscle weakness with no alternative explanation. There is no standard diagnostic protocol or specific diagnostic tools, and revised criteria of El Escorial (3), Awaji’s algorithm (3) and Gold Coast criteria (4) are mainly used for patients’ inclusion in clinical trials. The lack of consensus and tools causes a delay of up to one year in the diagnosis, which worsens the prognosis of the disease.

Only one drug has been approved by the European Medicines Agency against ALS, Riluzole, a glutamate antagonist that offers a marginal yet statistically significant extension of survival (5). Despite scientific advances, clinical trials for new drugs continue to face challenges because ALS is treated as a single disease without considering the cause or the mechanisms involved, which vary greatly depending on the case (1).

As with most neurodegenerative pathologies, ALS is believed to occur as a combination of ageing-associated dysfunctions, genetic predispositions and environmental factors. There is great pathological diversity across more than 20 associated genes. About 15% of cases stem from autosomal dominant factors due to mutations that affect protein degradation pathways and may favour the accumulation of TDP-43 (TBK1, OPTN, SQSTM1, UBQLN2, C9orf72 and VCP), metabolism pathways of RNA (TARDBP, FUS, MATR3, TIA1, hnRNPA1 and ATXN2), and dynamics of the cytoskeleton and axonal transport (TUBA4A, PFN1, KIF5A and DCTN1), among other genes involved (SOD1). Environmental risk factors include smoking, body mass index, physical exercise, occupational and environmental exposure to metals, pesticides, *β*-methylamino-l-alanine, head injuries, and viral infections. Despite these associations, the definitive causal relationship with ALS has not been established (1).

Clinically, the onset of ALS starts with weakness and focal muscle atrophy that subsequently spreads as the disease progresses. There is great variability regarding the initial site of manifestation (usually limbs), the age of onset (58-63 years for sALS and 40-60 years for fALS) and the rate of progression. The average survival period after the onset of symptoms is three years, where death is caused by respiratory failure associated with disease progression. Beyond motor difficulties, about 50% of patients suffer from extra motor manifestations, which are limited to mild behavioural and/or cognitive changes in 35-40% of cases. However, 10-15% experienced an additional diagnosis of frontotemporal dementia (FTD). This complex condition involves degeneration of frontal and anterior temporal lobes that causes behaviour changes, executive functioning impairment, and/or language impairment by molecular mechanisms underlying both FTD and ALS (1).

The pathogenic profile of ALS includes loss of neuromuscular connection, axonal retraction, and subsequent cell death of UMN and LMN, surrounded by astrogliosis and microgliosis, with ubiquitinated inclusions in surviving neurons with TDP-43 as the major component in more than 95% of ALS cases (6). Multiple molecular pathways are implicated in the pathogenesis, such as failures in proteostasis, neuroinflammation, excitotoxicity, mitochondrial dysfunction and oxidative stress, oligodendrocyte dysfunction, cytoskeletal abnormalities and axonal transport defects, RNA metabolism abnormalities, nucleocytoplasmic transport deficits, and impaired DNA repair (1; 7). In brief, ALS mainly affects the processes of protein control and degradation, RNA metabolism, and cytoskeletal and axonal transport, making the cytoplasmic aggregation of TDP-43 the prevailing neuropathological hallmark as it is present in more than 95% of patients (1; 6).

### 1.2. TDP-43

TDP-43 (TAR DNA-binding protein 43) is a 414 amino acid RNA-binding protein. Therefore, it is involved in multiple RNA biogenesis and processing steps, such as transcription, splicing, microRNA maturation, and RNA transport (8). TDP-43’s two RNA recognition motifs (RRM1 and RRM2) allow the binding to GU repeats predominantly located in long introns and at the 3’UTR end of the mRNA (8; 9; 10). Furthermore, TDP-43 regulates the splicing of many RNAs, both coding and non-coding, including mRNAs encoding proteins involved in neuronal survival and various proteins relevant to neurodegenerative diseases (11; 12). It also stabilises the mRNA by recruiting CNOT7/ CAF1 deadenylase to the 3’UTR end, leading to deadenylation of the poly(A) tail and thus shortening (13). It also regulates the processing of mitochondrial transcripts, so it is involved in the maintenance of mitochondrial homeostasis (14).

In response to oxidative stress, TDP-43 associates with ribosomes by forming stress granules that promote cell survival (15; 16). Likewise, TDP-43 participates in the formation and regeneration of skeletal muscle through the formation of cytoplasmic myo-granules and the binding to mRNA encoding sarcomeric proteins (17). On the other hand, TDP-43 regulates the expression of HDAC6, ATG7 and VCP in a PPIA/ CYPA-dependent manner (17) and downregulates CDK6 expression (18). In addition, TDP-43 contributes to the maintenance of circadian clock periodicity by stabilising CRY1 and CRY2 proteins in an FBXL3-dependent manner (19).

Considering its nuclear and cytoplasmic functions, TDP-43 can move between the nucleus and the cytoplasm, and the cell nucleus is its main location. In addition, there is a small concentration of TDP-43 located in the mitochondria, and in cases of oxidative stress, it is located in the stress granules. TDP-43 is encoded by the TARDBP gene, which has 18,185 base pairs and is located at 1p36.22 in the positive or sense strand of chromosome 1 between base pairs 11,012,344 and 11,030,528. It has 38 transcripts, and 5,371 single nucleotide polymorphisms (SNPs) that affect one or more of the protein’s functions have been identified (20).

### 1.3. The TDP-43 Proteinopathy

The TDP-43 proteinopathy is characterised by cytoplasmic aggregation and nuclear depletion. Its prevalence is observed in 95% of ALS cases, in 40-50% of FTD cases (21), in 20-50% of patients with Alzheimer’s disease (22) and in the limbic structures of most patients with predominantly limbic age-related TDP-43 encephalopathy (LATE) (23; 24). These findings substantiate the association of TDP-43 pathology with neurodegeneration, particularly when explored within the framework of ALS.

The reasons for suboptimal localisation can be multifaceted. Primarily, at least 48 pathogenic dominant variants of the TARDBP gene have been associated with ALS, most of them being nonsense mutations in the glycine-rich region of the carboxy-terminal region (25). In these cases, the causal participation of this protein can be established in the pathogenesis of the disease. However, there are TARDBP mutations in only 3% of fALS cases and 1.5% of sALS cases (24; 26), meaning a significant proportion of patients exhibit TDP-43 proteinopathy in the absence of TARDBP gene mutations, necessitating the involvement of other genetic factors. TDP-43 proteinopathy has already been associated with C9ORF72 expansions, VCP (Valosin-containing protein) and TBK1 mutations, and hnRNPA1 and hnRNPA2B mutations in multisystem proteinopathies (MSPs) (22; 24; 27; 28; 29).

However, although associated genes have been identified, the pathogenic mechanism leading from genetic mutation to the formation of cellular inclusion and the subsequent onset of ALS remains ununderstood. Three possible hypotheses that could give rise to this type of proteinopathy are discussed: the acquisition of a gain-of-function, the emergence of a novel deleterious function and the manifestation of a loss-of-function scenario (30).

Ensuring the optimal functioning of TDP-43 mandates a meticulous balance in its expression levels. This homeostasis depends on a process of autoregulation, nucleocytoplasmic transport, correct liquid-liquid phase separation (LLPS), correct mitochondrial function, autophagy and RNA binding, so any of these processes’ alteration can lead to TDP-43 proteinopathy (31). In addition to mislocalisation and aggregation, the positive and negative involvement of certain post-translational modifications (PTMs), such as ubiquitination, proteolytic cleavage, phosphorylation, and others less identified, such as acetylation, sumoylation, and disulfide bond formation should be considered (31). The pathological role of these MPTs has not been fully studied, with phosphorylation and ubiquitination being the most understood.

Clinically, TDP-43 proteinopathy impairs the motor system and exacerbates cognitive decline, which explains its association with the FTD that sometimes accompanies ALS. At the microscopic level, gliosis and microvacuoles accompany synaptic and neuronal loss (21). Four morphological patterns of TDP-43 inclusions have been established to classify four types of FTD: type A is characterised by having small intracytoplasmic inclusions in cortical layers and intranuclear inclusions in superficial cortical layers II and III; type B presents round neuronal inclusions in the cortex; type C has long neurites immunoreactive to TDP-43; and in type D, intranuclear inclusions and cytoplasmic inclusions are distinguished in neurons (21; 31). However, these morphological patterns have not been specifically associated with ALS. Instead, cytoplasmic aggregates are discussed in a general and less detailed manner.

The involvement of TDP-43 proteinopathies beyond neurodegeneration in myopathies has also been studied. TDP-43 aggregates have been found in inclusion body myopathies such as sporadic inclusion body myositis and inclusion body myopathy associated with PCV multisystem proteinopathy (32; 33). The role of TDP-43 in muscle physiology and related diseases continues to be investigated (17; 34).

Currently, there are monoclonal antibodies designed to detect phosphoserines 409 and 410 of the C-terminal region of TDP-43 (35) as phosphorylation of these residues is only found in patients with FTD or ALS and therefore can be considered a pathological marker of TDP-43 proteinopathy. However, this marker is only one of the many traits that can occur and cannot be used as a diagnostic tool for proteinopathy. For this reason, it is necessary to develop tests or diagnostic tools for TDP-43 proteinopathy that can be used for population screening (36; 37).

### 1.4. Deep Learning

Deep Learning (DL), in its supervised version, seeks to overcome challenges thanks to its ability to learn through training (38). These models can be applied to very diverse fields, including but not limited to healthcare (39). The foundational architecture of DL is based on artificial neural networks, a type of model comprising an assembly of interconnected elementary processing units known as neurons that generate a series of real-value outputs processed through non-linear activation functions to obtain an objective final result (outcome). While the training phase of a DL model demands substantial time investment due to its considerations of myriad parameters, it requires little time to run in testing time (40).

One crucial factor in the final performance of DL models is the quality and quantity of the data it learns from. This data can be sequential (with a relevant order), 2D (digital images that are numerical matrices, symbols, expressions or pixels arranged in rows and columns) or tabular (headed rows and columns organised as a dataset, following a logical and systematic arrangement of the data in the form of rows and columns that are based on the properties or characteristics of the data) (38). Furthermore, the models can be combined, forming complex architectures that are able to deal with more intricate data types, such as videos.

DL can be classified into three main categories according to the existence of labelled information (ground truth) that guides the training. These types are (1) supervised learning, where label data exists; (2) unsupervised learning, in which the absence of labelling data only allows the characterisation of high-order correlation properties and the analysis of patterns; and (3) semi-supervised learning, where label data is partial and requires a combination of both methods. The choice of a model will depend on the objective or problem to be solved and the availability of ground truth information (38).

Among the most widely used supervised learning models to handle 2D data are Convolutional Neural Networks (CNNs). Different CNN variants are used for visual recognition, medical image analysis, and image segmentation (38; 41). These models learn directly from the input without the need for human intervention to extract features, elaborating multiple convolutions and pooling layers so that each layer considers the optimal parameters to obtain a significant output and reduce the complexity of the model. In addition, overfitting, the problem of excessive adjustment to training data under limited sample sizes, can be ameliorated with tools like dropout, regularisation, early stopping and data augmentation (42).

### 1.5. Goals

We aimed to develop CNN models capable of detecting TDP-43 protein pathology in high-resolution microscopy images and to explore its relevance in distinguishing ALS from control samples based on post-mortem tissue. We used a dataset of high-resolution microscopy images provided by Dr Mathew Horrocks’ research group at the University of Edinburgh’s School of Chemistry. These images were obtained from post-mortem brain samples using immunofluorescence techniques to detect nuclei (DAPI) and super-resolution microscopy to highlight TDP-43 protein (pTDP-43^409/410^ and Apt-1) (43).

We subsequently processed these images to develop the CNN models based on the classification of samples by the presence or absence of ALS. However, initially, these models did not obtain substantial discriminative results. Our main objective of this paper is to understand and optimise the image data to support the model through a series of statistical analyses. By assessing the samples to be used for classification, we expect to identify any variable that may be useful for guiding the model. As a secondary objective, we seek to discern whether there are significant differences between the samples of the different degrees of protein pathology established by clinicians from the University of Edinburgh and assess the patient’s inter-variability.

Samples were analysed for five variables: cytoplasmic TDP-43 (red pixels), nuclear TDP-43 (pink pixels), the total amount of TDP-43 (red plus pink pixels), and cytoplasmic and nuclear ratios (red between red and pink sum or pink between red and pink sum respectively). These features were used to relate our samples with the existing bibliography, determine the most balanced classification, and understand variables with significant differences between groups. Problematic samples were detected and deleted to repeat the statistical analysis and generate more significant conclusions. Two new datasets were created, compared and tested. Finally, the reduced dataset was used as input to a hierarchical clustering algorithm using Euclidean distance as the distance metric. The resulting dendrogram can provide insights into the structure and relationships within the data and help tailor classification models using the generated clusters of the data. Ultimately, the aim is to improve the performance and interpretability of binary classification tasks.

## 2. Methodology

### 2.1. Samples

Statistical analyses were conducted on the image dataset provided by the University of Edinburgh for the development of our CNN models. These samples were sourced from the Edinburgh Brain and Tissue Bank (EBTB), established by the UK Medical Research Council and collected to facilitate the creation of a super-resolution microscopy-compatible probe aiming to characterise individual TDP-43 aggregates, resulting in the creation of Apt-1, an RNA aptamer capable of binding to the pathologic TDP-43 protein in vitro. Apt-1 demonstrated superior discriminatory capability between ALS cases and healthy controls in post-mortem samples compared to pTDP-43^409/410^. Furthermore, the extensively characterised primary antibody also exhibited binding affinity to pathologic TDP-43 (43).

Our study utilised post-mortem tissue samples from Brodmann area 4 (motor cortex) obtained from 48 individuals. The dataset supported three potential classification tasks: detection of TDP-43 pathology (binary), grading of pathology severity (multiclass), and ALS diagnosis vs. control (binary). However, only the first was selected for model development due to data limitations (see Methods for details). Among these, 31 were identified as having ALS based on clinical data, including genetic information and neuropsychological assessment using the ECAS (Edinburgh Cognitive and Behavioural ALS Screen). Each ALS sample was dewaxed and rehydrated to undergo immunohistochemistry (IHC) with the primary rabbit polyclonal antibody pTDP-43^409/410^ and staining with haematoxylin. These samples were recorded as Zeis.czi files using the fluorescence imaging mode of the ZEISS Axio Scan.Z1 scanner coupled to the brightfield microscope.

Ten areas of interest were sectioned from each sample using Napari to evaluate the number of neurons and chromogen-positive glial cells (Table 1). These cell-type-specific annotations were later used to assess intra-sample heterogeneity and inform the pathology grade assignment. Subsequently, the dataset passed a thresholding process with Python 3.9 and Fiji 1.53, resulting in images with a black background, green signal for the antibody, red for the aptamer, and blue for the cell nuclei (43).

**Table 1:**
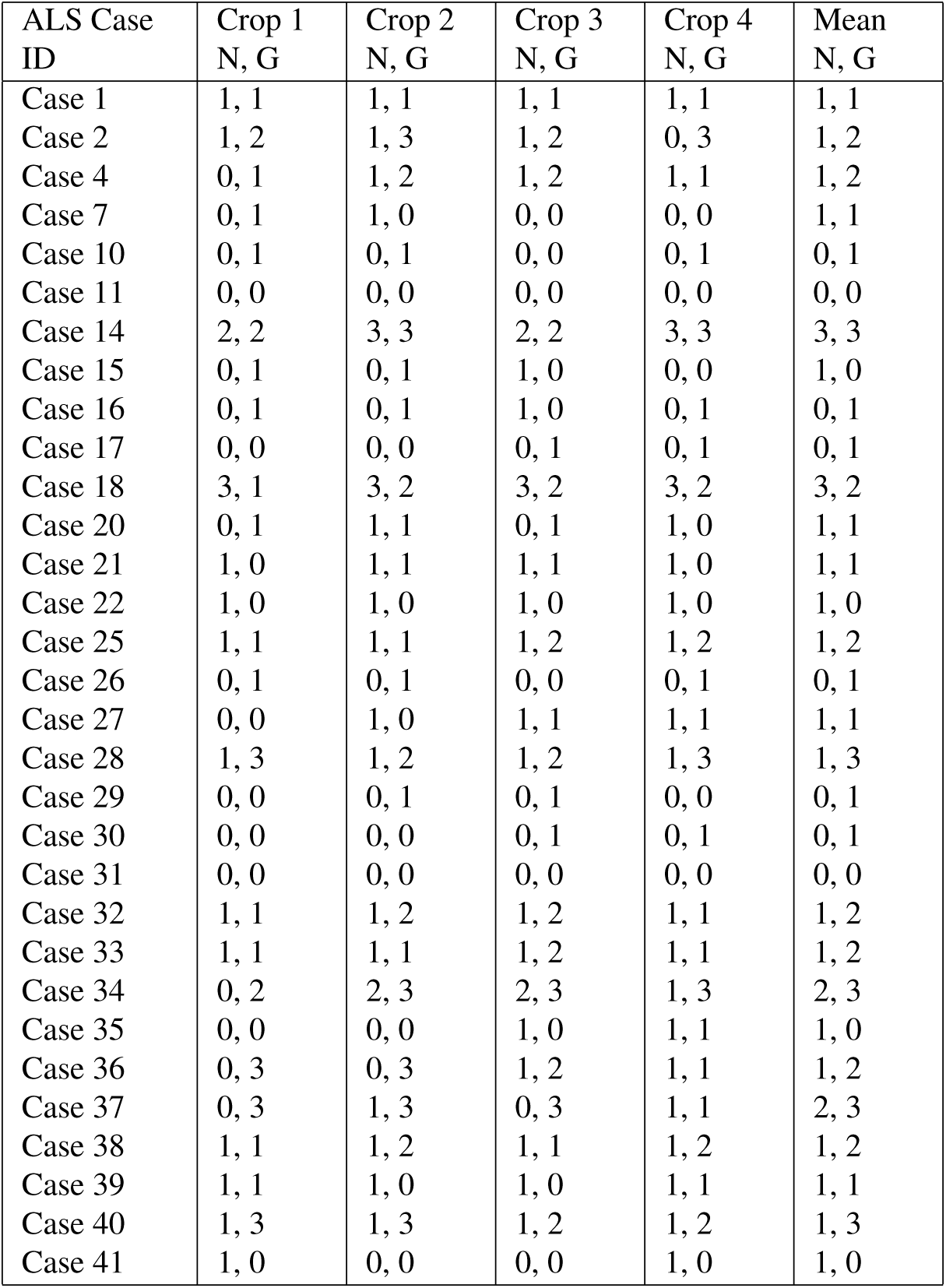
Grade of TDP-43 Pathology assigned to each area of interest for neurons (N), glial cells (G) and mean for each ALS sample (43).

| ALS Case ID | Crop 1<br>N, G | Crop 2<br>N, G | Crop 3<br>N, G | Crop 4<br>N, G | Mean<br>N, G |
| --- | --- | --- | --- | --- | --- |
| Case 1 | 1, 1 | 1, 1 | 1, 1 | 1, 1 | 1, 1 |
| Case 2 | 1, 2 | 1, 3 | 1, 2 | 0, 3 | 1, 2 |
| Case 4 | 0, 1 | 1, 2 | 1, 2 | 1, 1 | 1, 2 |
| Case 7 | 0, 1 | 1, 0 | 0, 0 | 0, 0 | 1, 1 |
| Case 10 | 0, 1 | 0, 1 | 0, 0 | 0, 1 | 0, 1 |
| Case 11 | 0, 0 | 0, 0 | 0, 0 | 0, 0 | 0, 0 |
| Case 14 | 2, 2 | 3, 3 | 2, 2 | 3, 3 | 3, 3 |
| Case 15 | 0, 1 | 0, 1 | 1, 0 | 0, 0 | 1, 0 |
| Case 16 | 0, 1 | 0, 1 | 1, 0 | 0, 1 | 0, 1 |
| Case 17 | 0, 0 | 0, 0 | 0, 1 | 0, 1 | 0, 1 |
| Case 18 | 3, 1 | 3, 2 | 3, 2 | 3, 2 | 3, 2 |
| Case 20 | 0, 1 | 1, 1 | 0, 1 | 1, 0 | 1, 1 |
| Case 21 | 1, 0 | 1, 1 | 1, 1 | 1, 0 | 1, 1 |
| Case 22 | 1, 0 | 1, 0 | 1, 0 | 1, 0 | 1, 0 |
| Case 25 | 1, 1 | 1, 1 | 1, 2 | 1, 2 | 1, 2 |
| Case 26 | 0, 1 | 0, 1 | 0, 0 | 0, 1 | 0, 1 |
| Case 27 | 0, 0 | 1, 0 | 1, 1 | 1, 1 | 1, 1 |
| Case 28 | 1, 3 | 1, 2 | 1, 2 | 1, 3 | 1, 3 |
| Case 29 | 0, 0 | 0, 1 | 0, 1 | 0, 0 | 0, 1 |
| Case 30 | 0, 0 | 0, 0 | 0, 1 | 0, 1 | 0, 1 |
| Case 31 | 0, 0 | 0, 0 | 0, 0 | 0, 0 | 0, 0 |
| Case 32 | 1, 1 | 1, 2 | 1, 2 | 1, 1 | 1, 2 |
| Case 33 | 1, 1 | 1, 1 | 1, 2 | 1, 1 | 1, 2 |
| Case 34 | 0, 2 | 2, 3 | 2, 3 | 1, 3 | 2, 3 |
| Case 35 | 0, 0 | 0, 0 | 1, 0 | 1, 1 | 1, 0 |
| Case 36 | 0, 3 | 0, 3 | 1, 2 | 1, 1 | 1, 2 |
| Case 37 | 0, 3 | 1, 3 | 0, 3 | 1, 1 | 2, 3 |
| Case 38 | 1, 1 | 1, 2 | 1, 1 | 1, 2 | 1, 2 |
| Case 39 | 1, 1 | 1, 0 | 1, 0 | 1, 1 | 1, 1 |
| Case 40 | 1, 3 | 1, 3 | 1, 2 | 1, 2 | 1, 3 |
| Case 41 | 1, 0 | 0, 0 | 0, 0 | 1, 0 | 1, 0 |

Based on this data, four grades of TDP-43 pathology were clinically assigned as follows: 0 (None or control), 1 (Mild), 2 (Moderate) and 3 (Severe), using the total number of chromogen-positive cells in the sample and the corresponding assigned grades (Table 2). However, these grades were binarised for training the classification model (presence vs. absence of pathology) due to class imbalance across categories.

**Table 2:** Number of TDP-43 positive cells in an area of interest related to each grade of TDP-43 pathology.

| Grade of TDP-43 Pathology | Number of TDP-43 positive cells in an area of interest |
| --- | --- |
| 0 (Control or None) | 0 |
| 1 (Mild) | 1-5 |
| 2 (Moderate) | 6-14 |
| 3 (Severe) | +15 |

For this study, we analysed images corresponding to Crop 1 and Crop 2 of each tissue sample, with ten regions of interest (ROIs) selected per crop, resulting in a total of twenty ROIs per subject. These were processed to eliminate the signal corresponding to pTDP-43^409/410^ and keep only the DAPI nuclear signal and Apt-1 signal due to the superior ability of Apt-1 versus pTDP-43^409/410^ to distinguish ALS cases and controls (43).

### 2.2. Ethics Statement

This study exclusively utilised pre-existing, anonymised image data generated from post-mortem tissue samples. No new data were collected, and no direct interaction with human participants occurred. The data were originally obtained from the Medical Research Council (MRC) Edinburgh Brain and Tissue Bank. Clinical information related to ALS cases was collected in vivo through the Scottish Motor Neuron Disease Register and the CARE-MND (Care Audit Research and Evaluation for Motor Neuron Disease) platform, with informed consent obtained from all participants. A psychological assessment using the ECAS was performed prior to death. Ethical approval and consent for tissue collection and data usage were secured by the original data providers in accordance with relevant guidelines. As this study involved secondary analysis of fully anonymised data, no additional ethical approval was required.

### 2.3. Data Augmentation

For the initial development of the deep learning models, the dataset underwent a data augmentation process to balance the number of healthy samples with those from ALS patients. This was done using non-destructive geometrical transformations to increase the training sample size and improve model generalisation.

To ensure a systematic and reproducible augmentation procedure, we used the Python Imaging Library (PIL) to generate 16 augmented versions of each original image. These were created by applying all combinations of vertical and horizontal flipping, followed by rotations of 90°, 180°, and 270°. This also included rotated versions of the flipped images, resulting in comprehensive augmentation coverage applied equally across all classes. Due to the initial class imbalance (i.e., more ALS than control samples), we subsequently applied random downsampling of ALS images post-augmentation to achieve class balance. The augmentation script is provided as Supplementary Code.

It is important to note that while the augmented dataset was used to train and evaluate the deep learning models, the original (non-augmented) image dataset was retained for all statistical analyses. This ensured that the statistical characterisation and class comparisons were based solely on the true, unmodified data.

### 2.4. Feature-Based Dataset for Statistical Analysis

From the original annotations and image metadata, a structured dataset was created in Microsoft Excel, compiling relevant information such as patient ID, sex, age, tissue section (1–2), image number (1–10), TDP-43 pathology classification (Yes/No), TDP-43 pathology grade (0–3), and quantitative measurements including red and pink pixel counts, total TDP-43 signal, cytoplasmic and nuclear proportions, and their ratios. These variables served as extracted features used to construct a secondary dataset for statistical analysis.

Three classification problems were considered: (1) the presence or absence of TDP-43 pathology, where any section graded as 0 (including both healthy controls and ALS cases without detectable TDP-43 pathology) was considered as absence of pathology; (2) the severity of TDP-43 pathology based on the provided clinical grading. The grading system establishes five groups: 0-Control (healthy controls), 0-No (ALS cases without detectable TDP-43 pathology), 1-Mild, 2-Moderate, and 3-Severe, based on the number of TDP-43-positive cells observed. Finally, (3) classification of ALS diagnosis, where samples listed in Table 1 are considered pathological (ALS), and the remaining are controls.

Then, the number of red and pink pixels in each sample was calculated and included as features in the dataset. Red pixels indicate the signal emitted by Apt-1, meaning the presence of TDP-43. Pink pixels are due to the combination of the blue nuclear signal (DAPI) and red pixels (Apt-1). Therefore, red pixels represent cytoplasmic TDP-43, while pink pixels indicate nuclear TDP-43. A Python self-made code generated these numerical data from the images to understand the relationship of the samples with the existing literature on TDP-43 proteinopathy. The ratios of red and pink pixel counts were calculated, along with their sum, representing the total amount of TDP-43. From this, the proportion of cytoplasmic TDP-43 (red pixels divided by the total of red and pink pixels) and nuclear TDP-43 (pink pixels divided by the same total) were computed, as well as the ratio of cytoplasmic to nuclear signal.

### 2.5. Deep Learning Model Architecture and Training

We evaluated the performance of multiple convolutional neural network (CNN) architectures for classifying highresolution microscopy images as either TDP-43 proteinopathic or non-pathological. Standard architectures such as VGG16 and ResNet50 were used, along with a custom-designed CNN model implemented in Keras.

The VGG16 architecture accepts input images of size 224 × 224 × 3 and passes them through a series of convolutional layers with 3 × 3 filters and a stride of 1. Five max-pooling layers (2 × 2) are interleaved throughout the network. Feature extraction is followed by three fully connected layers (4,096, 4,096, and 1,000 neurons, respectively), with ReLU activation for all intermediate layers and a final sigmoid activation for binary classification. Designed to overcome limitations in training deep networks, ResNet50 uses residual blocks and shortcut connections. The architecture begins with a 7 × 7 convolutional layer (stride 2) and a 3 × 3 max-pooling layer, followed by several convolutional blocks with residual connections. The network ends with an average pooling layer and a fully connected output layer with a sigmoid activation.

The detailed layer structures of the ResNet50 and VGG16 architectures used in this study are illustrated in Figure 1. All models were compiled using the binary cross-entropy loss function and optimised with the Adam optimiser using a learning rate of 0.001. Training was conducted over 40 epochs with a batch size of 32. To prevent overfitting, early stopping was applied based on validation accuracy, with a patience of one epoch.

**Figure 1:**
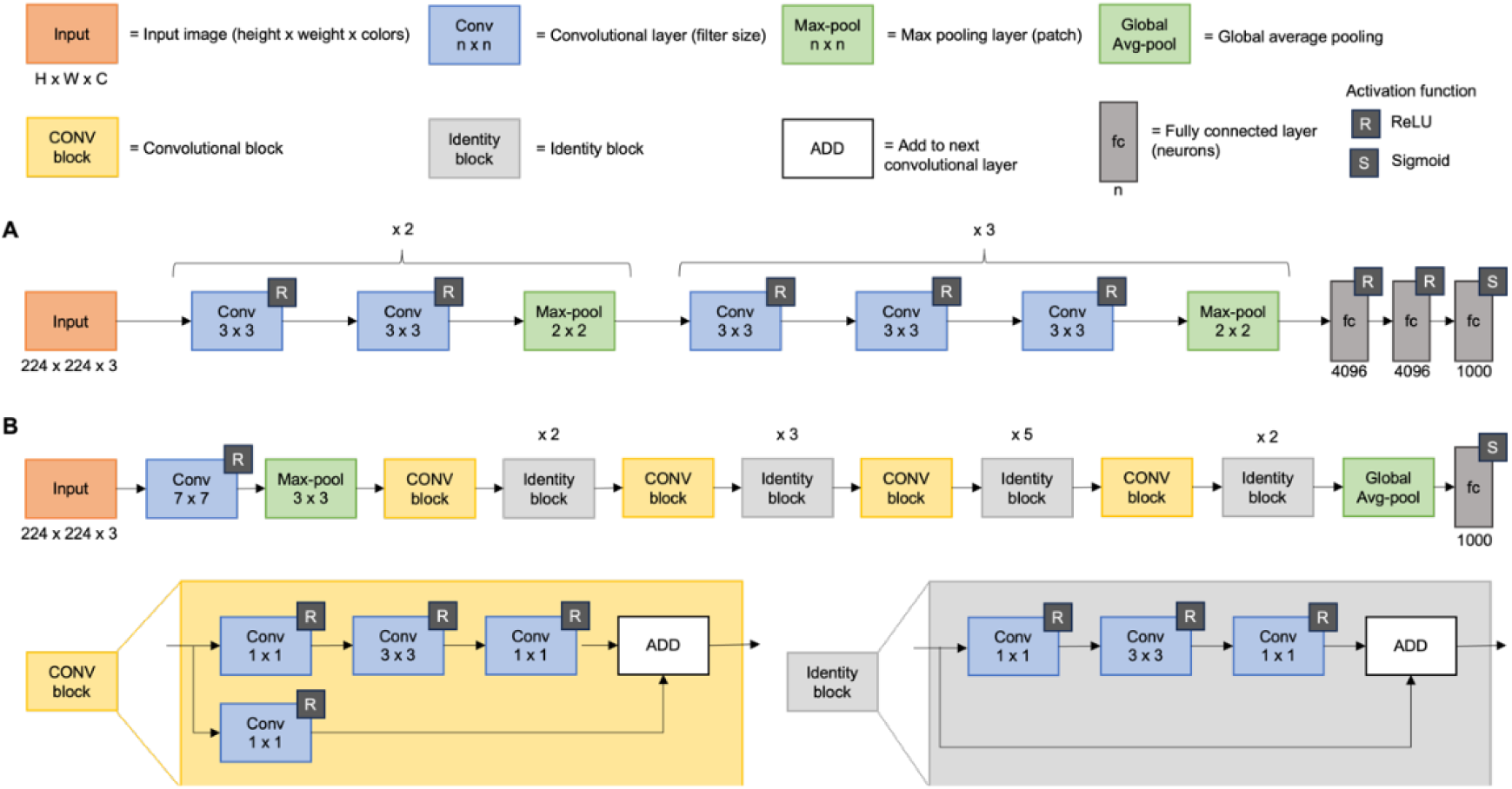
Architectural overview of the convolutional neural networks used in the study: VGG16 (A) and ResNet50 (B).

### 2.6. Initial Analysis

Descriptive analysis was carried out using XLSTAT Software (version 2023.3) and Microsoft Excel (version 2021), focusing on three classification categories: TDP-43 Pathology, TDP-43 Pathology Grades, and ALS. The analysis aimed to evaluate pixel frequency distributions and determine the most balanced and viable classification sets for training the deep learning models. Balanced sample classes were prioritised to prevent potential generalisation issues, which could lead the model to predominantly classify samples into the majority group (44).

Subsequently, samples for the three classification problems were analysed based on three variables: (1) the total amount of TDP-43 (measured by the sum of red and pink pixels), (2) cytoplasmic proportion (ratio of red pixels to total pixels), and (3) nuclear proportion (ratio of pink pixels to total pixels). Sampling distributions were visually compared between groups using scattergrams.

The statistical significance of differences between groups for the three variables was evaluated using Student’s t-tests for binary classifications (presence/absence of TDP-43 pathology and ALS) and ANOVA for the classification by grades of TDP-43 pathology. Pairwise comparisons among all grades were performed using Student’s t-tests to elucidate the differences.

### 2.7. Dataset Curation

Given the inconclusive results obtained in the initial statistical analysis (see Section 3.1), it was necessary to clean the dataset and repeat the analysis to clarify or confirm different conclusions. We observed that the sampling distribution for the cytoplasmic TDP-43 proportion tended to cluster around values of 0 and 1, which suggested potential issues during image acquisition or processing (Figures B.12, B.13, and B.15).

The criteria for sample exclusion focused on the cytoplasmic TDP-43 proportions. Samples with values near 0 or 1 were considered problematic. Specifically, values approaching 1 may reflect insufficient colocalisation between DAPI and Apt-1 signals, potentially due to imaging artefacts or technical variability. Conversely, values near 0 could suggest binding issues, possibly related to aptamer degradation or inconsistencies in the staining protocol. The threshold for exclusion was set at cytoplasmic ratios less than 0.1 or greater than 0.9, based on visual inspection and expert judgment.

A total of 297 samples were excluded based on the above criteria. Each excluded sample was documented and annotated (Table 3) to track potential patterns of errors. We hypothesised that the majority of these samples were affected by isolated errors during image acquisition or processing. Approximately 70% of the excluded samples exhibited cytoplasmic TDP-43 ratios close to 1, suggesting systematic issues in the colocalisation protocol involving Apt-1 and DAPI.

**Table 3:** Deleted samples in the dataset curation process per subject and their characteristics (ALS, TDP-43 and grade).

| Deleted Samples |  |  |  |  |
| --- | --- | --- | --- | --- |
| Patient | Deleted Samples | ALS | TDP-43 | Grade |
| Case 1 | 1.1; 1.2; 1.3; 1.4; 1.5; 1.6; 1.7; 1.8; 1.9; 1.10; 2.1; 2.2; 2.4; 2.5 | Yes | Yes | 1-Mild |
| Case 2 | 1.1; 1.2; 1.3; 1.4; 1.5; 1.6; 1.7; 1.8; 1.9; 1.10; 2.1; 2.2; 2.3; 2.4; 2.5; 2.6; 2.7; 2.8; 2.9; 2.10 | Yes | Yes | 2-Moderate |
| Case 3 | 1.2; 1.5; 2.5 | No | No | 0-Control |
| Case 4 | 1.8 | Yes | Yes | 2-Moderate |
| Case 5 | 1.1; 1.2; 1.3; 1.10; 2.4; 2.5 | No | No | 0-Control |
| Case 6 | 1.6; 1.8; 2.2; 2.4; 2.6; 2.7; 2.8; 2.10 | No | No | 0-Control |
| Case 7 | 1.1; 1.2; 1.3; 1.4; 1.5; 1.6; 1.7; 1.8; 1.9; 1.10; 2.1; 2.2; 2.3; 2.4; 2.5; 2.6; 2.7; 2.8; 2.9; 2.10 | Yes | Yes | 1-Mild |
| Case 8 | 1.1; 1.7; 2.8 | No | No | 0-Control |
| Case 9 | 2.1 | No | No | 0-Control |
| Case 10 | 1.1; 1.2; 1.3; 1.4; 1.5; 1.6; 1.7; 1.8; 1.9; 1.10; 2.1; 2.2; 2.3; 2.4; 2.5; 2.6; 2.7; 2.8; 2.9; 2.10 | Yes | Yes | 1-Mild |
| Case 11 | 1.1; 1.4; 1.5; 1.6; 1.7; 1.8; 1.9; 1.10; 1.11; 2.1; 2.2; 2.3; 2.4; 2.5; 2.6; 2.7; 2.8; 2.9; 2.10 | Yes | No | 0-None |
| Case 12 | 1.1; 1.2; 1.3; 1.4; 1.5; 1.6; 2.3; 2.5; 2.8; 2.10 | No | No | 0-Control |
| Case 13 | 1.7; 2.2; 2.5 | No | No | 0-Control |
| Case 14 | 1.8 | Yes | Yes | 3-Severe |
| Case 15 | 1.5 | Yes | Yes | 1-Mild |
| Case 16 | 1.1; 1.2; 1.3; 1.4; 1.5; 1.6; 1.7; 1.8; 1.9; 1.10; 2.1; 2.2; 2.3; 2.4; 2.5; 2.6; 2.7; 2.8; 2.9; 2.10 | Yes | Yes | 1-Mild |
| Case 17 | 1.3; 1.5; 1.6; 1.7; 1.8; 1.9; 1.10; 2.1; 2.3; 2.4; 2.5; 2.7; 2.8; 2.10 | Yes | No | 1-Mild |
| Case 18 | 1.2; 1.4; 1.5; 1.6; 1.10; 2.6; 2.7; 2.8; 2.9; 2.10 | Yes | Yes | 3-Severe |
| Case 19 | 2.2; 2.10 | No | No | 0-Control |
| Case 20 | 1.1; 1.3; 1.4; 1.6; 1.9 | Yes | Yes | 1-Mild |
| Case 21 | 1.8; 2.6; 2.8; 2.9; 2.10 | Yes | Yes | 1-Mild |
| Case 22 | 1.1; 1.3; 1.4; 1.7; 2.3 | Yes | Yes | 1-Mild |
| Case 23 | 2.1; 2.2; 2.4; 2.5; 2.7; 2.8; 2.10 | No | No | 0-Control |
| Case 24 | 2.1; 2.4; 2.8; 2.10 | No | No | 0-Control |
| Case 25 | 1.1; 1.2; 1.3; 1.4; 1.5; 1.6; 1.7; 1.8; 1.9; 1.10; 2.1; 2.2; 2.3; 2.4; 2.5; 2.6; 2.7; 2.8; 2.9 | Yes | Yes | 2-Moderate |
| Case 26 | 2.1; 2.2; 2.4; 2.5; 2.6; 2.7, 2.8 | Yes | Yes | 1-Mild |
| Case 27 | 1.1; 1.3; 1.4; 1.5; 1.6; 1.7; 1.8; 1.9; 2.1; 2.2; 2.3; 2.5; 2.7; 2.8 | Yes | No | 1-Mild |
| Case 28 | 1.1; 1.2; 1.3; 1.4; 1.5; 1.6; 1.7; 1.8; 1.9; 1.10; 2.1; 2.2; 2.5; 2.6; 2.8; 2.9 | Yes | Yes | 3-Severe |
| Case 29 | 1.2; 1.4; 1.5; 1.6; 1.7; 1.9; 2.8 | Yes | No | 1-Mild |
| Case 30 | 1.1; 1.2; 1.3; 1.4; 1.7; 1.10; 2.1; 2.2; 2.3; 2.4; 2.6; 2.7; 2.9; 2.10 | Yes | No | 1-Mild |
| Case 31 | 1.2; 1.3; 1.4; 1.5; 1.9; 1.10; 2.1; 2.2; 2.3; 2.4; 2.5; 2.6; 2.7; 2.8; 2.9; 2.10 | Yes | No | 0-None |

After curation, the dataset became more balanced, with fewer extreme outliers. This adjustment was expected to improve the robustness of the subsequent statistical analysis and model training. The revised dataset was subjected to the same statistical tests as the initial dataset, including t-tests for binary comparisons and ANOVA for multiple groups. Post-hoc pairwise comparisons were also repeated to confirm the presence of statistically significant differences.

Following dataset curation, the results were reevaluated to determine whether the cleaned dataset yielded clearer and more reliable conclusions.

### 2.8. Final Analysis

The statistical analysis conducted on the initial dataset was repeated for the curated dataset. Frequencies were analysed to assess the most balanced classification sample sets. Scattergrams were used to compare the sampling distribution between groups for each classification scenario, focusing on three variables: total amount of TDP-43, cytoplasmic proportion, and nuclear proportion.

The existence of statistically significant differences between groups for the three variables was assessed using Student’s t-tests for binary classifications, such as TDP-43 pathology presence and ALS diagnosis. For these analyses, the assumption of normality was verified using the Shapiro-Wilk test. Given that some groups did not follow a normal distribution, Mann-Whitney tests were applied to evaluate differences across five variables: the number of red and pink pixels (cytoplasmic and nuclear TDP-43, respectively), total TDP-43 signal, and the cytoplasmic and nuclear proportions of TDP-43.

Additionally, differences between grades of TDP-43 pathology were evaluated using ANOVA. Pairwise comparisons between grades were also performed using independent t-tests to understand the specific differences among the categories. These steps were taken to ensure robust analysis and to identify statistically significant patterns within the dataset following curation.

### 2.9. Datasets’ Testing

Both the initial and curated datasets were compared to examine the sampling distribution across patients. For this purpose, box plots were generated based on five variables: the number of red and pink pixels (representing cytoplasmic and nuclear TDP-43, respectively), the total amount of TDP-43 (sum of red and pink pixels), and the proportions of cytoplasmic and nuclear TDP-43.

To evaluate classification performance, we created two datasets distinguishing control samples from TDP-43 pathological samples, using both the initial and curated versions. For each classification task, the samples were stratified and randomly divided into training (80%) and testing (20%) sets, maintaining class balance within each split.

Given the limited number of patients (48 in total), we implemented 15 independent, stratified train-test splits to assess model robustness and reduce dependency on any single data partition. This approach enabled us to obtain a distribution of performance metrics, with final results reported as the average across all repetitions. While K-fold cross-validation is a commonly used strategy for small datasets, we opted for repeated stratified splits to better assess variability under different partitionings. This method has been adopted in several medical imaging studies facing similar challenges related to dataset size and class imbalance (45; 46; 47).

To provide a comprehensive assessment of classification performance, the following metrics were computed for both training and validation sets: accuracy, sensitivity, specificity, F1-score, and Matthews Correlation Coefficient (MCC). Values were recorded for each run and averaged to report the final results.

For the curated dataset, the mean and standard deviation of each of the five variables were calculated per patient. This analysis aimed to capture inter-subject variability, providing insight into how individual differences may affect model performance. Additionally, we explored subject-level variability using hierarchical clustering, where all samples from each individual were grouped together. The resulting dendrogram provided a visual representation of relationships within the dataset and highlighted potential issues in class distribution, offering a deeper understanding of the dataset’s structure post-curation.

## 3. Results

### 3.1. Initial Analysis

First, a descriptive analysis of the initial dataset containing all the available samples was performed. Groups’ frequencies were extracted from this analysis (Figure (2–1)). According to the classification by TDP-43 proteinopathy (See Figure (2-1A), 479 samples presented pathology (53%) compared to 420 healthy samples (47%). In the ALS classification (Figure (2-1B), 609 samples (68%) showed pathology compared to 290 healthy samples (32%). For the classification by grades of TDP-43 (Figure (2-1C) pathology, 290 controls (32%), 41 cases without protein pathology (5%), 309 mild cases (34%), 139 moderate cases (15%) and 120 severe cases (13%) were registered.

In the same way, it was possible to compare the sampling distribution for the cytoplasmic and nuclear proportion variables with the use of scattergrams for the classifications by TDP-43 pathology, see Figure B.12, depicting grades of TDP-43 pathology in Figure B.14, and distinguishing ALS in Figure B.13.

Student’s t-tests were performed with a 95% confidence interval (*α* = 0.05), and p-values below this threshold were considered statistically significant. The smaller the p-value relative to *α*, the stronger the evidence against the null hypothesis. For the classification by TDP-43 pathology (Figure (3–1)), significant differences were detected for the total amount of TDP-43 (p=0.003), with pathological samples having more protein. Significant differences were also detected for the cytoplasmic proportion (p<0.0001), which turned out to be lower for the pathological samples but not for the nuclear proportion (p=0.266).

For the classification by ALS (Figure (3–1)), no significant differences were detected for the total amount of TDP-43 (p=0.216). However, significant differences were found for the cytoplasmic (p<0.0001) and nuclear (p= 0.046) proportions, as pathological samples have smaller values for both variables.

Afterwards, ANOVA was used with a confidence interval of 95% for comparing the different protein pathology grades. It was determined that there were significant differences between groups for the three variables (total amount of TDP-43 and the cytoplasmic and nuclear ratios) as p<0.0001. In addition, it was possible to graphically compare the differences of the means for the three variables (Figure B.9).

Additionally, grades of TDP-43 pathology were compared independently with Student’s t-tests. The results were grouped into tables, assessing the level of significance of each comparison (Figure 4).

**Figure 2:**
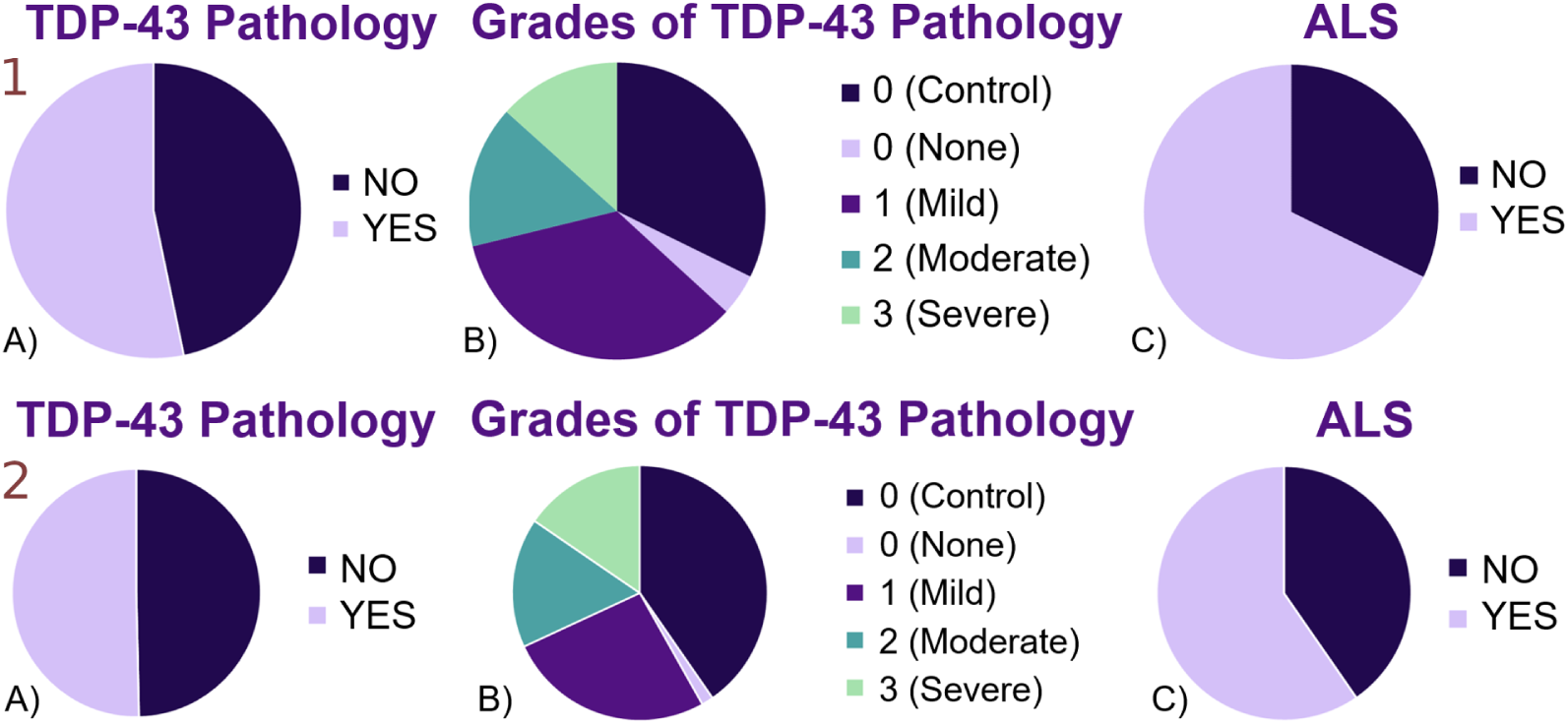
Frequency graphs of the initial (1) and final (2) complete dataset for the classifications of (A) TDP-43 Pathology, (B) TDP-43 Pathology Grades, and (C) ALS.

**Figure 3:**
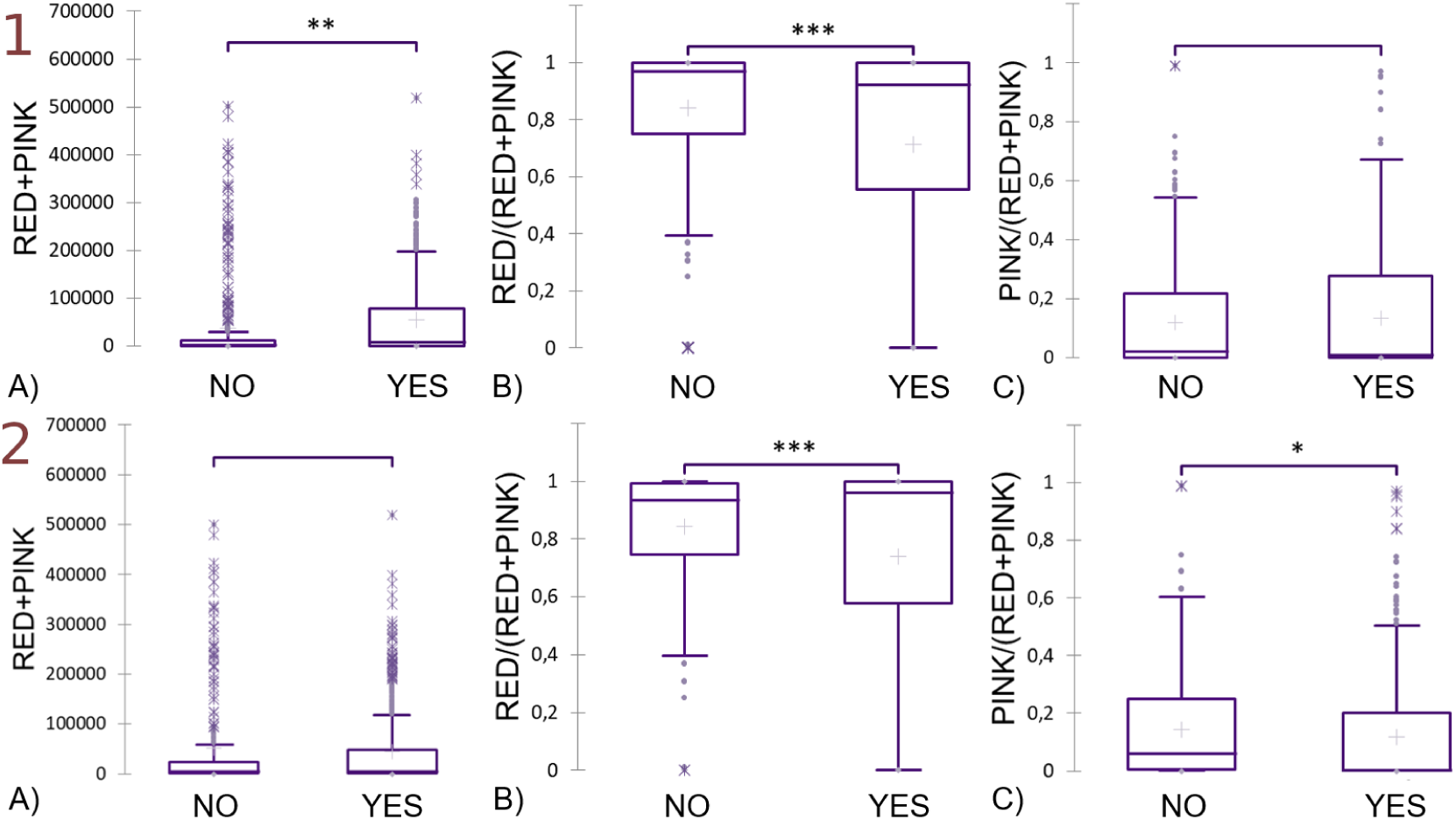
(1) Comparative plots of means for the. (A) total amount of TDP-43, (B) cytoplasmic proportion, and (C) nuclear proportion for the classification of TDP-43 pathology using the complete dataset. (2) Comparative plots of means for the same variables in the classification of ALS. Statistical significance was assessed using Student’s t-test (*α* = 0.05), with significance levels denoted as follows: (*) p < 0.05, (**) p < 0.01, (***) p < 0.001.

**Figure 4:**
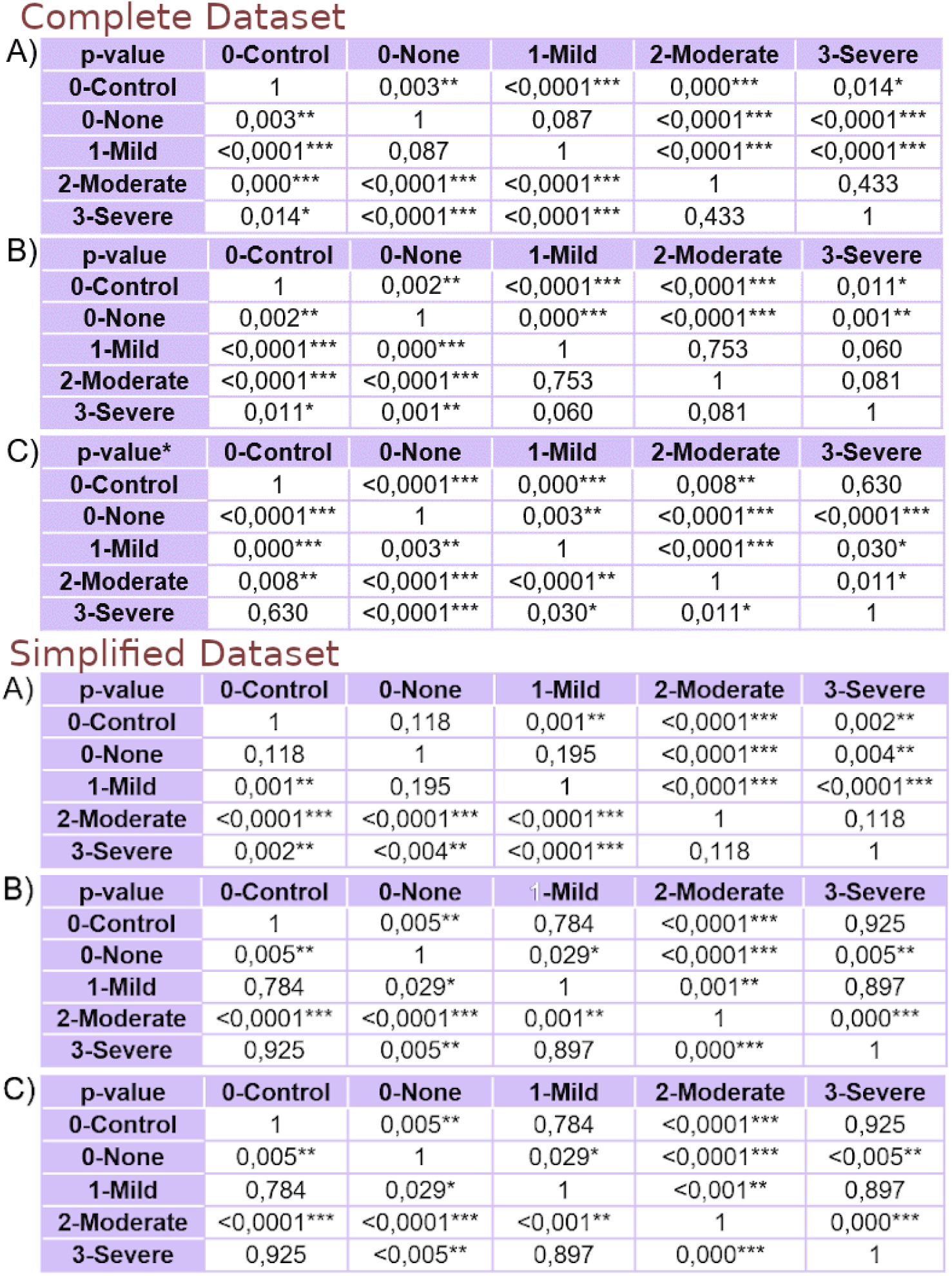
Complete Dataset – P-values comparing the means of the different TDP-43 pathology grades for. (A) total amount of TDP-43, (B) cytoplasmic proportion, and (C) nuclear proportion. **Simplified Dataset** – P-values for the same comparisons using the simplified dataset. Statistical significance was assessed using Student’s t-test (*α* = 0.05), with significance levels indicated as follows: (*) p < 0.05, (**) p < 0.01, (***) p < 0.001.

### 3.2. Final Analysis

The dataset was cleaned up based on the previous results and the same procedures were repeated on this new reduced dataset. Initially, multiple descriptive analyses were performed, from which groups’ frequencies were extracted (Figure (2–2)). According to the classification by TDP-43 pathology, 303 samples presented pathology (50.33%) compared to the 299 healthy cases (49.67%). In the ALS classification, 359 samples (60%) showed pathology compared to 243 healthy ones (40%). For the classification by grades of TDP-43 pathology, 243 controls (40%), 9 cases without protein pathology (1%), 158 mild cases (26%), 99 moderate cases (16%) and 93 severe cases (15%) were registered. In the same way, the sampling distribution for the cytoplasmic and nuclear proportion variables was compared for the classifications by TDP-43 pathology (Figure B.16), by grades of TDP-43 pathology (Figure B.15) and by ALS (Figure B.17).

Afterwards, differences between groups were verified using Student’s t-tests with *α*=0.05 so that any p-value lower than *α* was considered statistically significant. The significance level is greater as the p-value decreases with respect to *α*. For the classification by ALS (Figure (5–1)), no significant differences were detected neither for the total amount of TDP-43 (p=0.137) nor for the cytoplasmic (p=0.107) or nuclear (p=0.107) proportions. On the other hand, for the classification by TDP-43 pathology (Figure (5–2)), significant differences were detected for the total amount of TDP-43 (p<0.0001), the cytoplasmic proportion (p<0.009), and the nuclear proportion (p=0.009). In this case, pathological samples showed a lower cytoplasmic TDP-43 proportion and higher values for the nuclear proportion and total TDP-43.

In order to better understand these results for the classification by TDP-43 pathology, significant differences for both cytoplasmic (p=0,001) and nuclear (p<0,0001) concentrations of TDP-43 (red and pink pixels, respectively) were found according to Student’s t-tests (Figure (5–3)), having pathological samples higher values for both variables. For this dataset, the outcomes from ANOVA determined that there were significant differences between groups for the three variables since p<0.0001 for all three variables. In addition, it was possible to graphically compare the differences of the means for the three variables (Figure B.10). Severe and moderate TDP-43 pathological samples have higher total amounts of TDP-43 than controls. Severe samples show more cytoplasmic TDP-43 and less nuclear TDP-43 than moderate samples.

The different grades of TDP-43 pathology were compared independently using Student’s t-tests. Results were grouped in tables and analysed by assessing the levels of significance of each classification problem (Figure 4).

Finally, since the initial analysis showed that the samples did not follow a normal distribution, verified by the Saphiro-Wilk tests for all groups, differences for classification by TDP-43 pathology were verified with Mann-Whitney tests (Figure (5–4). Pathological samples revealed higher values for the total amount of protein (p<0,0001), the nuclear (p=0,038) proportion and nuclear (p<0,0001) and cytoplasmic (p<0,0001) amounts of TDP-43, despite a lower cytoplasmic proportion (p=0,038).

### 3.3. Datasets’ Testing

The distribution of each patient’s values for each variable was compared between the initial and the reduced datasets (Figure B.11). Afterwards, a ResNet-50 model (48) was trained and tested 15 times to assess the robustness and variability of the results with the two datasets.Violin plots were generated to visualise the distribution of performance metrics—accuracy, specificity, sensitivity, Matthews correlation coefficient (49), and F1-score—across 15 independent runs for both training and testing datasets (Figure 6).

**Figure 5:**
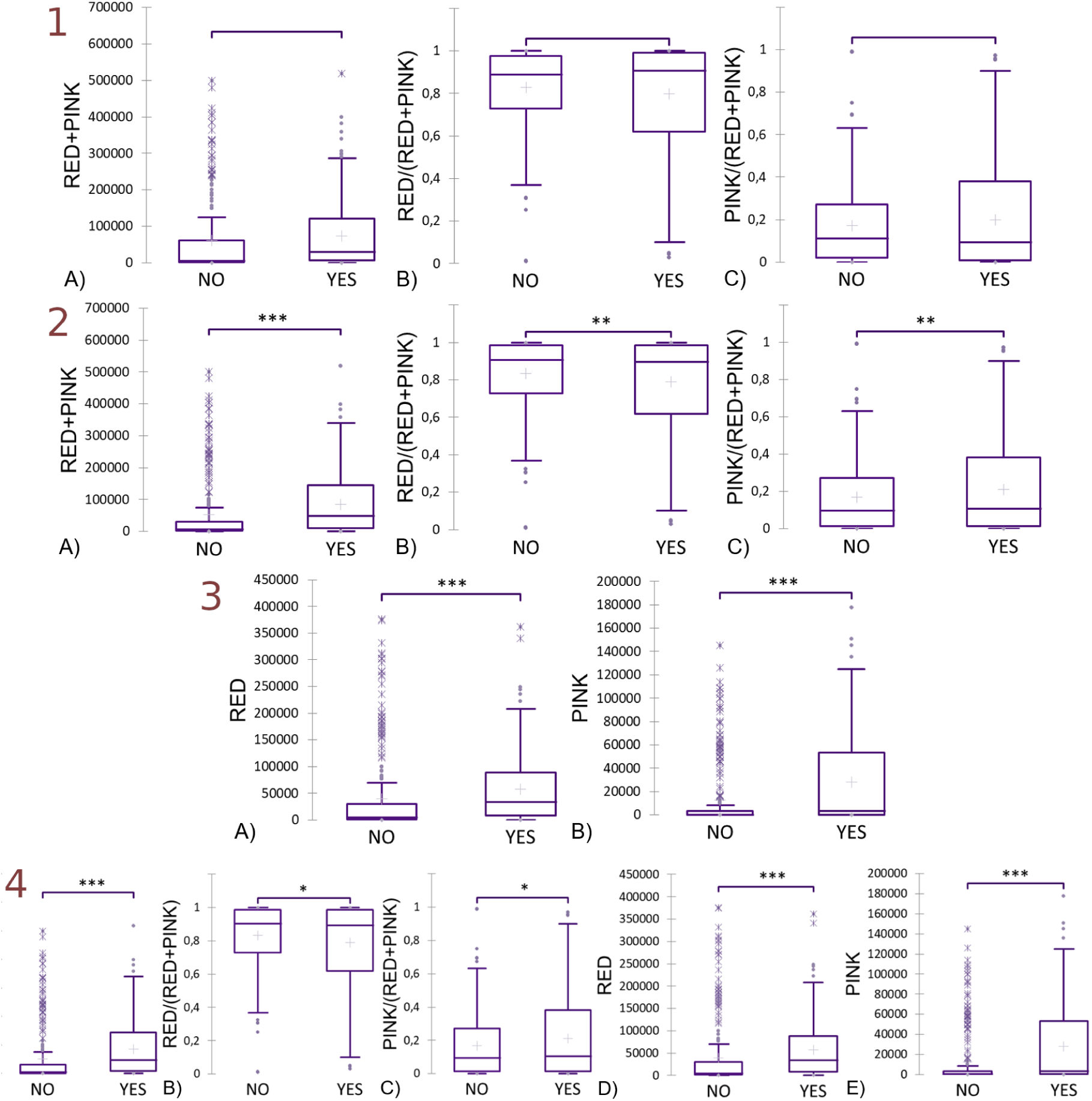
(1) Comparative plots of means for. (A) total TDP-43 amount, (B) cytoplasmic proportion, and (C) nuclear proportion in the ALS classification of the simplified dataset. (**2**) Comparative plots of the same variables in the classification of TDP-43 pathology using the simplified dataset. (**3**) Comparative plots of means for the (A) cytoplasmic and (B) nuclear TDP-43 amounts in the classification of TDP-43 pathology. (**4**) Comparative plots of means for (A) total TDP-43 amount, (B) cytoplasmic proportion, (C) nuclear proportion, and (D) cytoplasmic and (E) nuclear TDP-43 amounts in the classification of TDP-43 pathology. Statistical significance was assessed using Student’s t-test (*α* = 0.05) for analyses (1–3) and the Mann–Whitney U test (*α* = 0.05) for analysis (4). Significance levels are indicated as follows: (*) p < 0.05, (**) p < 0.01, (***) p < 0.001.

**Figure 6:**
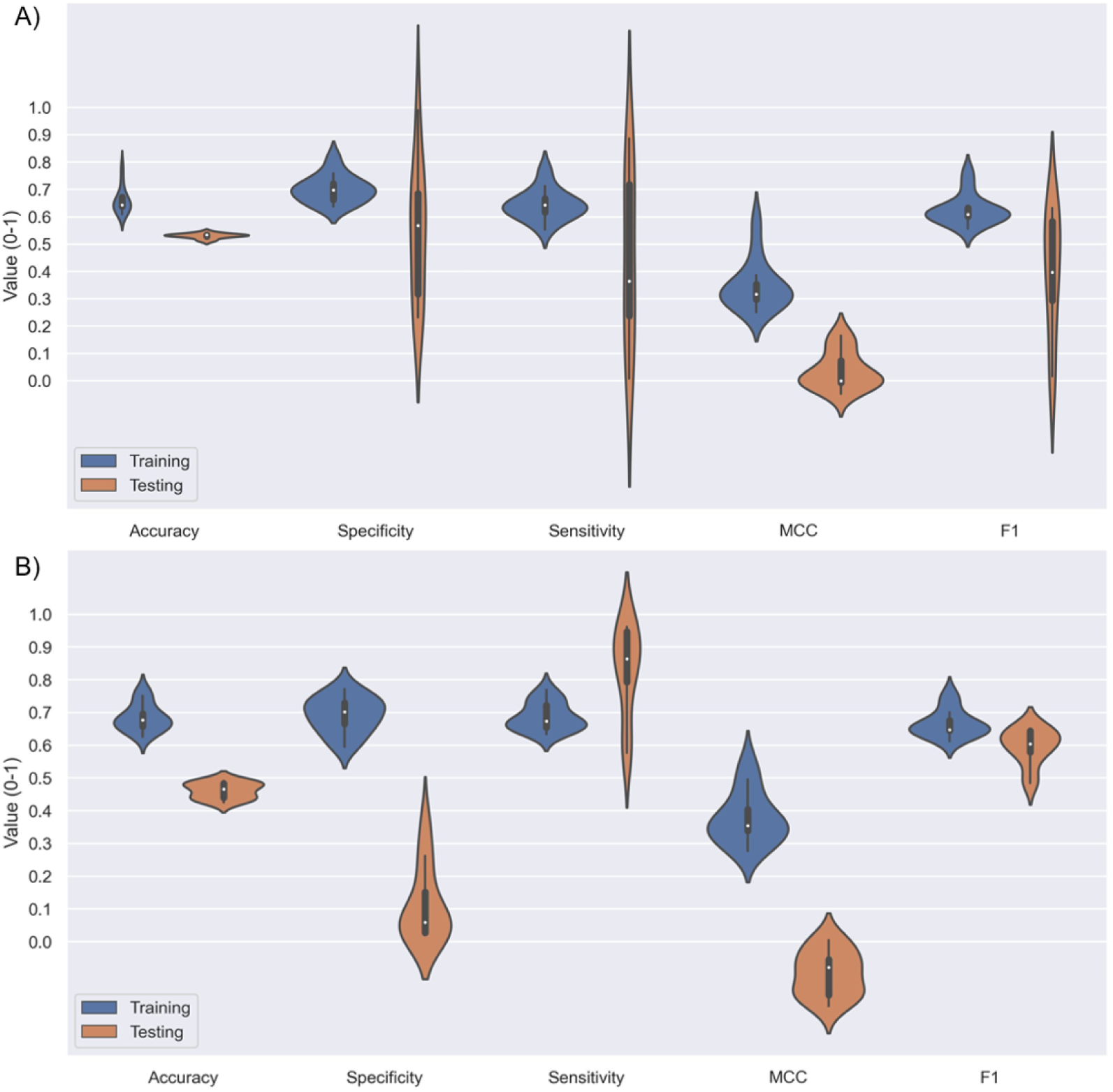
Violin plots for accuracy, specificity, sensitivity, Matthew’s correlation coefficient and F1-score resulting from training and testing the CNN model 15 times with (A) the complete and (B) the reduced datasets.

The dendrogram resulting from the hierarchical clustering of the reduced dataset was linked to the subject information to improve the understanding (Figure 7). Even if the dendrogram did not show the full distinction between groups, there are some trends that can be mentioned. The dendrogram showed two main clusters, separating 30 (on the left) from the other 11 subjects (on the right). The first group formed a three-subcluster division that was able to discriminate a first group with ALS and no TDP-43 proteinopathy, a second group with ALS and TDP-43 and a third group with no ALS and no proteinopathy. The second group on the right showed a much clearer structure, with a subgroup with no ALS/TDP-43 and a group with severe/moderate levels of the disease.

**Figure 7:**
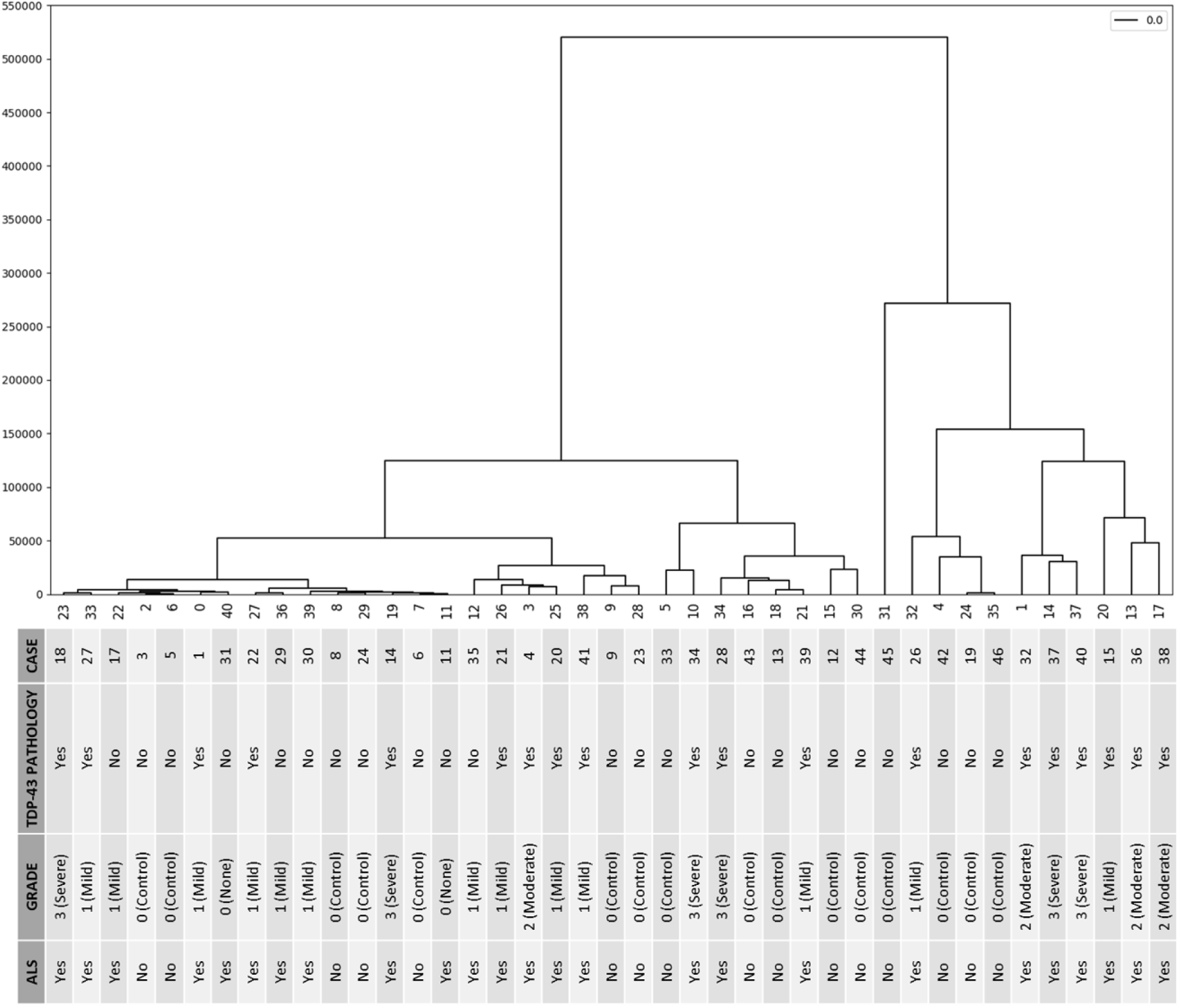
Hierarchical clustering graph between patients in the reduced dataset.

## 4. Discussion

The initial frequency analysis of sample types demonstrated a strong lack of balance between cases and controls for both ALS classification and pathology classification of TDP-43 (Figure (2–1)). This fact justifies the need for a process of oversampling the underrepresented group prior to training the models to balance the sample sets, followed by a general process of data augmentation to increase the total number of images and reach a higher generality. In addition, a clear correlation is observed between the samples with grade 0 of TDP-43 pathology and the control subjects of the ALS, noting that the final clinical diagnosis assigned is independent of the individual grades of each image sample. However, control samples used for ALS classification do not necessarily correspond to those used for TDP-43 pathology classification, as the latter is based on the grade assigned to each individual tissue section (Table 1), rather than on the overall patient diagnosis. This per-section grading captures intra-patient variability and provides a more objective basis for training, whereas assigning a single label per patient may introduce generalisation errors. It is more objective to consider each section’s grade separately and, therefore, the existent intra-patient variability. When assigning a unique classification label to all samples from a patient, a higher risk of generalisation can occur.

At this point, certain limitations related to the sample set must be considered. First, the initial sample size is limited, as often happens when dealing with low-incidence diseases. Secondly, samples of ALS controls were not clinically assessed for aggregated TDP-43, which can lead to certain biases by assuming that the controls do not have protein pathology. It is known that this proteinopathy may exist in the absence of ALS as it is compatible with other pathologies. On the other hand, the assignment of discrete grades (non, mild, moderate, and severe) is problematic. The initial annotation process associated the number of chromogen-positive cells to the assigned grade (Table 2). However, the grades are not equal (the number of cells does not increase proportionally), and it is not explained why those grades are established with those assignments (43). Furthermore, the number of chromogen-positive cells was not provided or published. Hence, we could not use it in this study to establish a more equitable association with the grades.

Samples with TDP-43 pathology were shown to have a superior total TDP-43 than controls but a lower proportion of protein in the cytoplasm (Figure (3–2)), suggesting that there is indeed an increased proportion of protein located in the nucleus although it is not significant, so results are not entirely conclusive. Considering these relevant results would be contradictory since TDP-43 proteinopathy has been characterised by a pattern of cytoplasmic aggregation and nuclear depletion, and the results obtained describe an opposite pattern.

For ALS classification, lower proportions of cytoplasmic and nuclear TDP-43 are observed in pathological samples without significantly altering the total amount of protein (Figure (5–1)). It could be said that these results are not conclusive as they are significant when considering only part of the information, but a minimum level of significance is not reached when considering all the information.

For the classification by grades of TDP-43 pathology, significant differences were determined for the three variables (total amount of TDP-43, cytoplasmic proportion, and nuclear proportion) according to the ANOVA results. If we compare the means of the two groups with grade 0, it is observed that controls have more total protein and higher nuclear proportion, while ALS samples have less total protein but a higher cytoplasmic proportion and a lower nuclear ratio (Figure B.10). When comparing the grades of pathology, an increase in the three variables is observed in grade 2 compared to grade 1, while these decrease in grade 3 with the exception of the cytoplasmic proportion, which keeps rising (Figure B.10). Therefore, differences will depend on which variable we measure and which grades we compare, varying the significance of these differences in each case (Figure 4).

Due to the lack of conclusive results to guide the DL model, it was proposed to reduce the dataset by removing problematic samples, being aware that this action would also harden the generalisation capability of the models. Sampling distribution of the complete dataset was analysed for the nuclear and cytoplasmic TDP-43 proportions variables for the three classifications (Figures B.12, B.14 and B.13), and a greater accumulation of samples was observed at values 0 and 1. Since at both physiological and pathological levels, there must be protein in both the nucleus and cytoplasm, it can be interpreted that samples that apparently do not show protein or only have it in one location are due to errors in materials or obtaining protocols and, therefore, do not represent reality. Thanks to this idea, the dataset was reduced by deleting these samples.

On the reduced dataset, an almost perfect balance was observed between samples with TDP-43 pathology and controls, but not for ALS (Figure (2–2)). The balanced dataset aimed at the classification of TDP-43 pathology is more optimal for training the DL models. As for the initial analysis, controls for the classifications by grades and by ALS are correlated because they share criteria, which is not the case for the classification by protein pathology, so the samples’ limitations and the criteria differences must always be considered.

When visualising the sampling distribution of the images from this reduced dataset for the cytoplasmic and nuclear proportions (Figures B.16, B.15 and B.17), a greater TDP-43 cytoplasmic proportion is still observed in all groups. Samples tend to group around values very close to 0 and 1, but since they are real values calculated based on the pixels, further reducing the dataset could cause the loss of valid information.

For the classification by TDP-43 pathology of this reduced dataset, significant differences were established for the three variables (Figure (5–2)) as controls had a higher proportion of cytoplasmic TDP-43 and, therefore, a lower proportion of nuclear protein, as well as less protein. This suggests that pathological samples generally have an excess of protein, so even with a lower cytoplasmic proportion, there would still generally be a greater amount of protein in the cytoplasm. Differences for the amounts of cytoplasmic and nuclear TDP-43 (red and pink pixels respectively) were analysed again with Student’s t-tests (Figure (5–3)) in order to verify that both amounts of TDP-43 were higher for pathological samples.

Samples with TDP-43 pathology had a general excess of protein with a significant increase in both cytoplasmic and nuclear protein concentration. This characteristic corroborates the literature in terms of the cytoplasmic TDP-43 aggregation pattern and the pathological profile of TDP-43 proteinopathy as the cytoplasmic levels of protein increase. However, cytoplasmic TDP-43 proportion decreases in pathological samples (1; 6). Nevertheless, it is impossible to report nuclear depletion as both the amount and proportion of nuclear TDP-43 increase in pathological samples.

As samples do not follow a normal distribution, which can be verified by Saphiro-Wilk tests for all groups, these differences were verified with Mann-Whitney tests (Figure 5-4). TDP-43 pathological samples showed a generalised protein excess with a significant increase of both cytoplasmic and nuclear amounts of protein, verifying the pattern of cytoplasmic aggregation (1; 6) with no nuclear depletion, although there is no increase of cytoplasmic proportion. Results for the classification by ALS (Figure (5–1)) determined that there are no differences between controls and pathological samples for any of the variables analysed, which would partially explain the initial results of the deep learning model based on this dataset.

Considering these previous points, it could be said that training the model based on the classification by pathology of TDP-43 could be more promising. In addition, the possibility of including some of the analysed variables in the current or new model could be studied in order to guide the model to the protein locations in the images.

Finally, for the classification by grades of TDP-43, significant differences were detected for the three variables. As for the complete dataset, the means of the groups vary in one way or another depending on the variable and the groups that are compared (Figure B.10), and in the same way, the significance of these differences varies according to the grades that are compared and the variable that is analysed in each case (Figure 4).

The dataset clean-up process deleted most of the outliers of every patient for the five analysed variables (Figure B.11). It also allowed us to see a more homogeneous pattern for cytoplasmic and nuclear proportions variables. Training with the reduced dataset generated slightly higher metric values for accuracy, specificity, sensitivity, Matthew’s correlation coefficient and F1-score. Testing the model with this dataset got more consistent results, although there is still a margin for improvement.

Regarding the dendrogram, it can be observed that there is no clear distinction between groups of any classification according to the clusters (Figure 7). This fact verifies that it is hard to distinguish samples according to analysed features and that the number of samples for some categories is limited.

Finally, in this study, we tested a limited number of CNN architectures and selected ResNet-50 based on its relatively better performance and stability. While Vision Transformers (50) (ViTs) are gaining popularity, they are less suitable for our setting due to the small dataset size and the lack of strong spatial dependencies in our microscopy images. ViTs typically require large-scale datasets and are designed to capture long-range patterns, which may not align well with the independent pixel structure of our data. Although we did not evaluate lightweight models such as EfficientNet (51) or Swin-Tiny (52) in this work, we recognise their potential and plan to investigate them in future studies.

## 5. Conclusion and Future Work

In conclusion, the analysis of the information from all the samples was inconclusive in guiding the DL model, but it facilitated the dataset-cleaning process. Based on the reduced dataset, the sample sizes for each class in the classification of TDP-43 pathology were closely matched, providing an ideal scenario for model training. In addition, there were significant differences between cases and controls for the total amount of TDP-43, its cytoplasmic and nuclear amounts and its cytoplasmic and nuclear proportions.

In this way, it was possible to guide the DL model based on the classification by TDP-43 pathology by ruling out the classification by ALS as a possibility. A new model possibility is proposed, considering variables for which cases and controls have significant differences. On the other hand, significant differences between the varied grades of protein pathology for the three variables have been clarified for both datasets. However, it must be understood that there is some variability in terms of these differences when individualising them, so that for each comparison between two groups, more or less significant differences will be found depending on the variable that is used. The reduced dataset has fewer outliers and resulted in better and more consistent results when training the model than the complete dataset, so the cleanup process was useful and efficient. However, the results were still insufficient as the clustering process did not group samples consistently. High-quality and more abundant samples are needed to produce better results. Thus, the goal of developing an effective TDP-43 pathology detection tool remains to be met.

In future work, we plan to incorporate K-fold cross-validation to complement the current evaluation strategy. This will provide an additional layer of validation and help ensure that every sample contributes to both training and testing phases. We also agree that exploring the remaining classification tasks (ALS vs. control and pathology grading) could strengthen the study, and we plan to revisit them as more annotated data become available. We will also explore t-SNE for feature space visualisation and alternative CAM-based methods, such as Score-CAM and SmoothGrad, to improve interpretability with high-resolution data.

Although the deep learning model did not achieve strong predictive performance, this outcome highlights a key insight of our study: the limitations in data quality, sample size, and annotation reliability significantly hinder model success. This finding underscores the importance of critically assessing dataset suitability before model deployment and offers a valuable reference for similar future studies in neuropathology.

## Data Availability

All data produced in the present study are available upon reasonable request to the authors.

## Acknowledgments

We would like to thank Dr Mathew Horrocks from the University of Edinburgh for providing the data used in this study. Their contributions were invaluable to the completion of this research.

## Author contributions statement

**Azucena Muñoz**: conceived the experiments, conducted the statistical experiments, wrote the first draft, and analysed the results. **Vasco Oliveira**: performed data curation and conducted the deep learning experiments. **Marta Vallejo:** provided resources, project administration, supervision, methodology, validation and content for revision versions. All authors reviewed the manuscript.

## Appendix A. Class Activation Maps for High-Resolution Images

Figure A.8 illustrates examples of Grad-CAM visualisations applied to high-resolution images. Although the heatmaps highlight the regions that contributed most to the model’s prediction, the spatial resolution of these overlays appears coarse relative to the fine-grained structure of the original data. This suggests that standard CAM methods may have limited interpretability when applied to super-resolution images. Future work will investigate alternative techniques, such as Score-CAM and SmoothGrad, which may offer better spatial localisation and interpretability in this context.

**Figure A.8:**
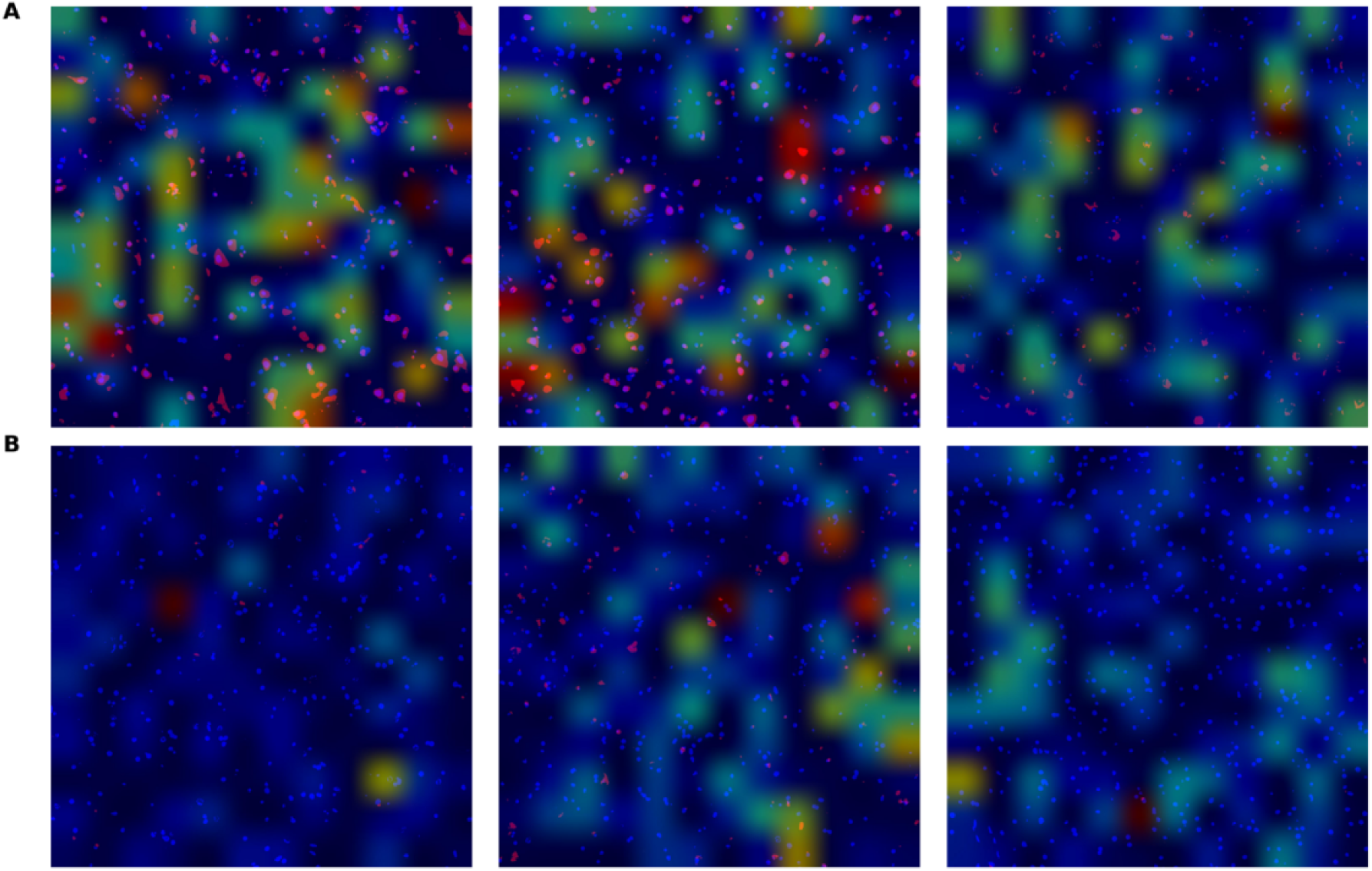
Representative Grad-CAM overlays on high-resolution microscopy images. Panel A shows three examples from TDP-43 positive samples, and Panel B shows three examples from control samples. The coloured heatmaps indicate the regions influencing the model’s predictions, with warmer colours (red/yellow) signifying stronger activation.

## Appendix B. Comparative graphs

Figure B.9 and B.10 show comparative graphs of the means of the different grades of TDP-43 pathology from the simplified dataset for the total amount of TDP-43.

Figure B.9 represents the results from the initial dataset, and Figure B.10 when the dataset was processed.

**Figure B.9:**
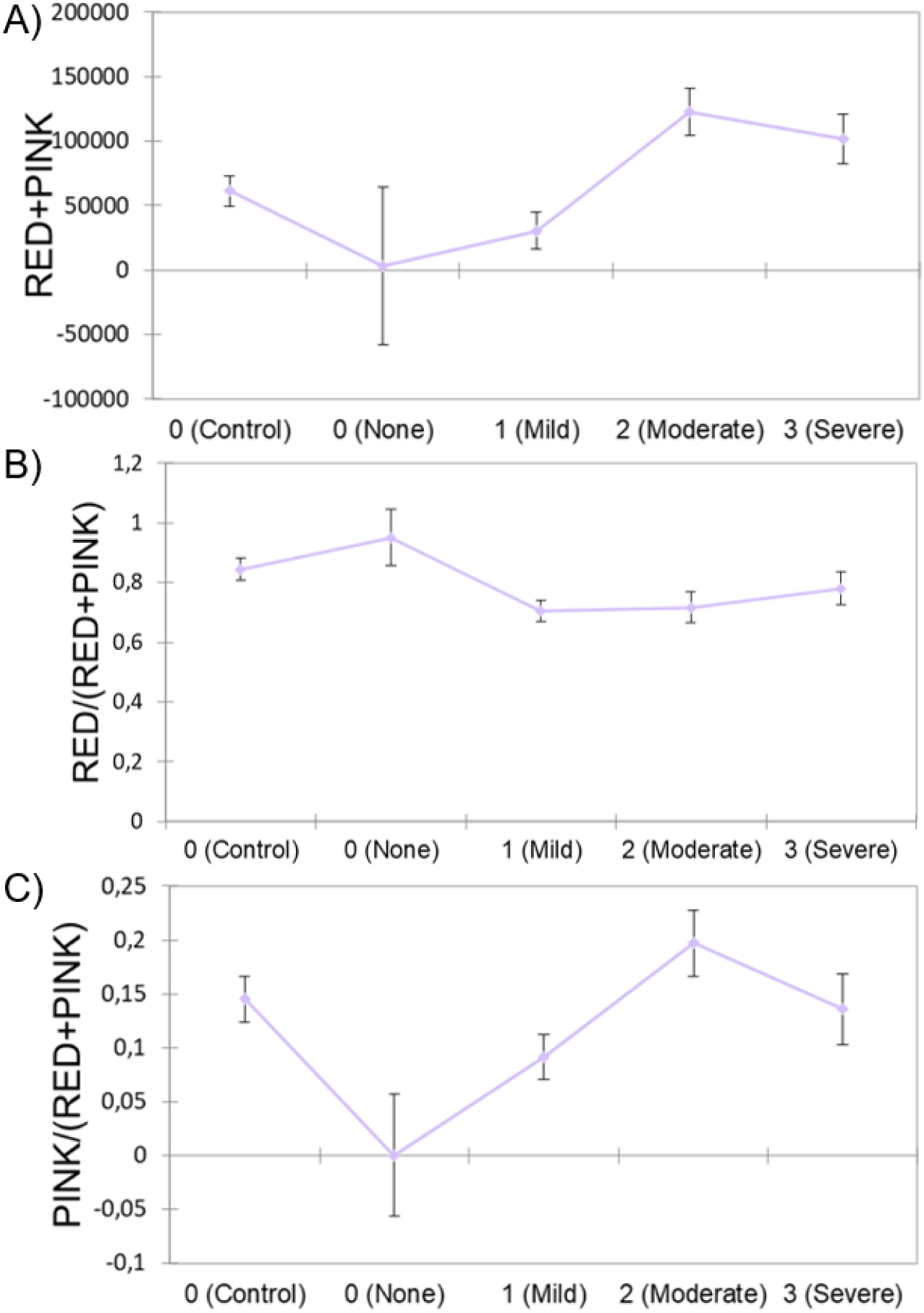
Comparative graphs of the means of the different grades of TDP-43 pathology from the complete dataset for the. (A) total amount of TDP-43 and the (B) cytoplasmic and (C) nuclear ratio.

**Figure B.10:**
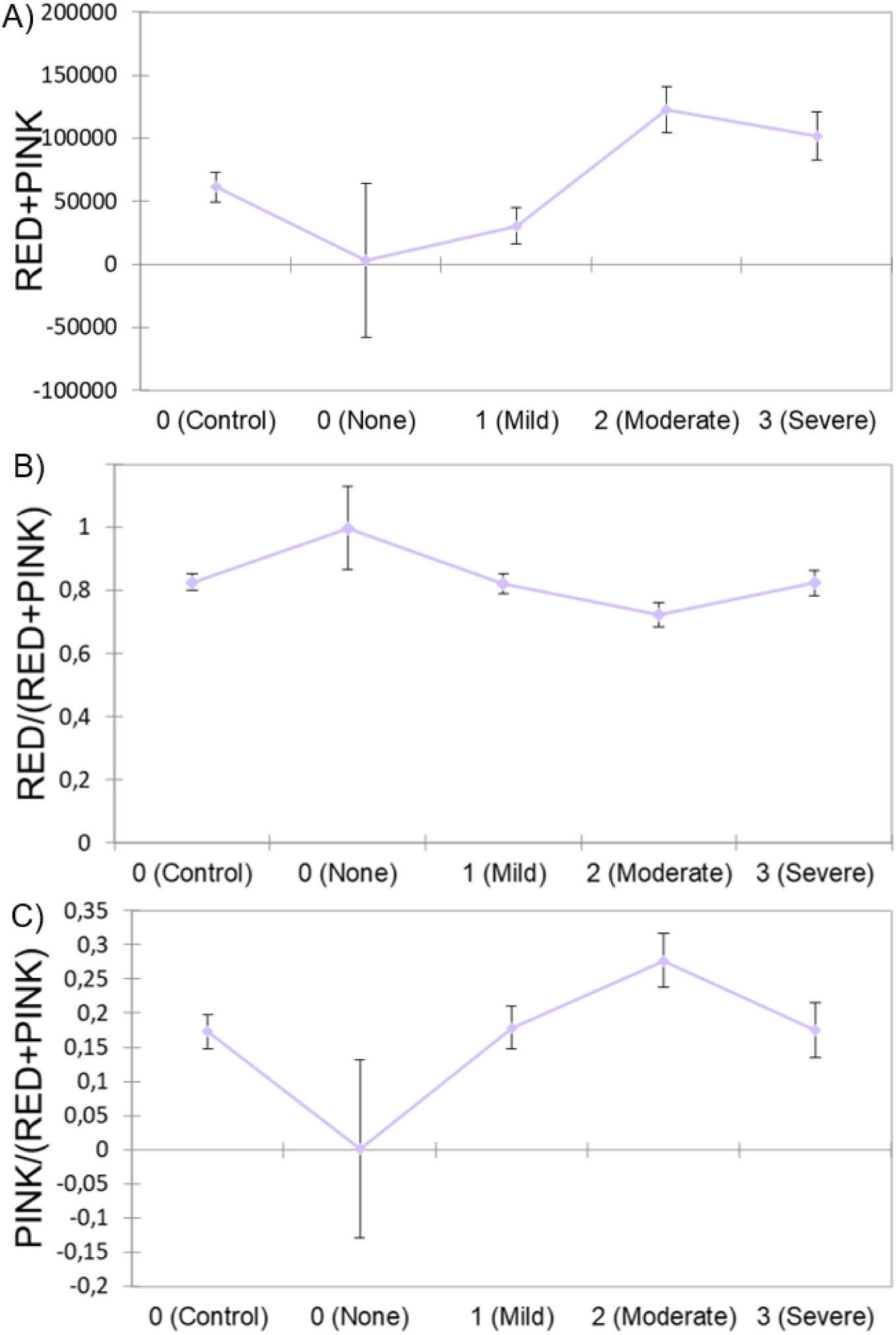
Comparative graphs of the means of the different grades of TDP-43 pathology from the simplified dataset for the. (A) total amount of TDP-43 and the (B) cytoplasmic and (C) nuclear proportions.

Figure B.11 depicts the comparison between the initial and the reduced dataset for five variables: the amount of cytoplasmic TDP-43 (red pixels), the amount of nuclear TDP-43 (pink pixels), the total amount of TDP-43 (red+pink) and cytoplasmic and nuclear ratios.

**Figure B.11:**
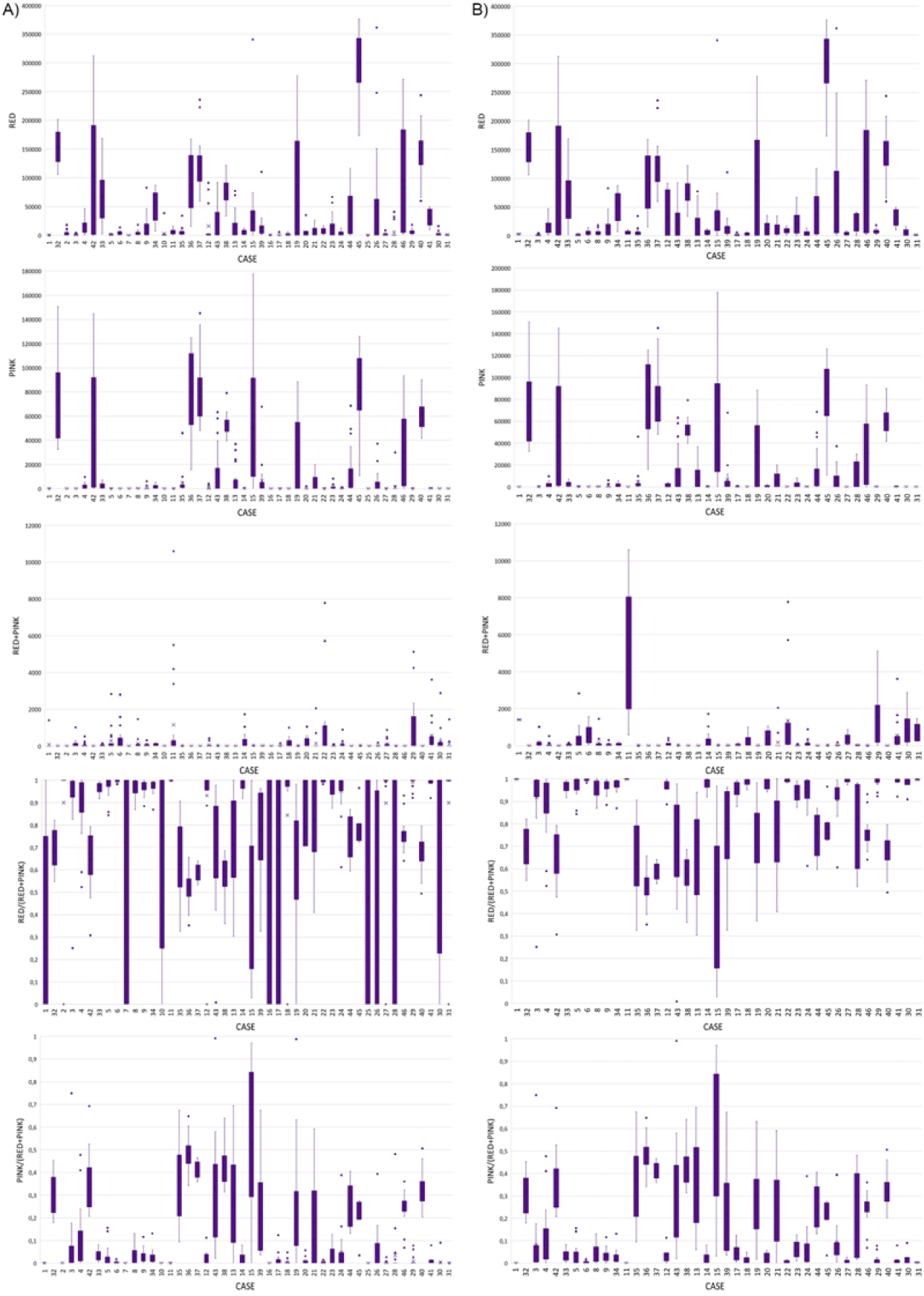
Comparison of (A) the initial dataset and (B) the reduced dataset for five variables: the amount of cytoplasmic TDP-43 (red pixels), amount of nuclear TDP-43 (pink pixels), the total amount of TDP-43 (red+pink) and cytoplasmic and nuclear ratios (red between red and pink sum or pink between red and pink sum respectively).

Figure B.12 represents the classification by TDP-43 pathology from the complete dataset for the variables cytoplasmic and nuclear proportion, and Figure B.13 for the classification by ALS.

**Figure B.12:**
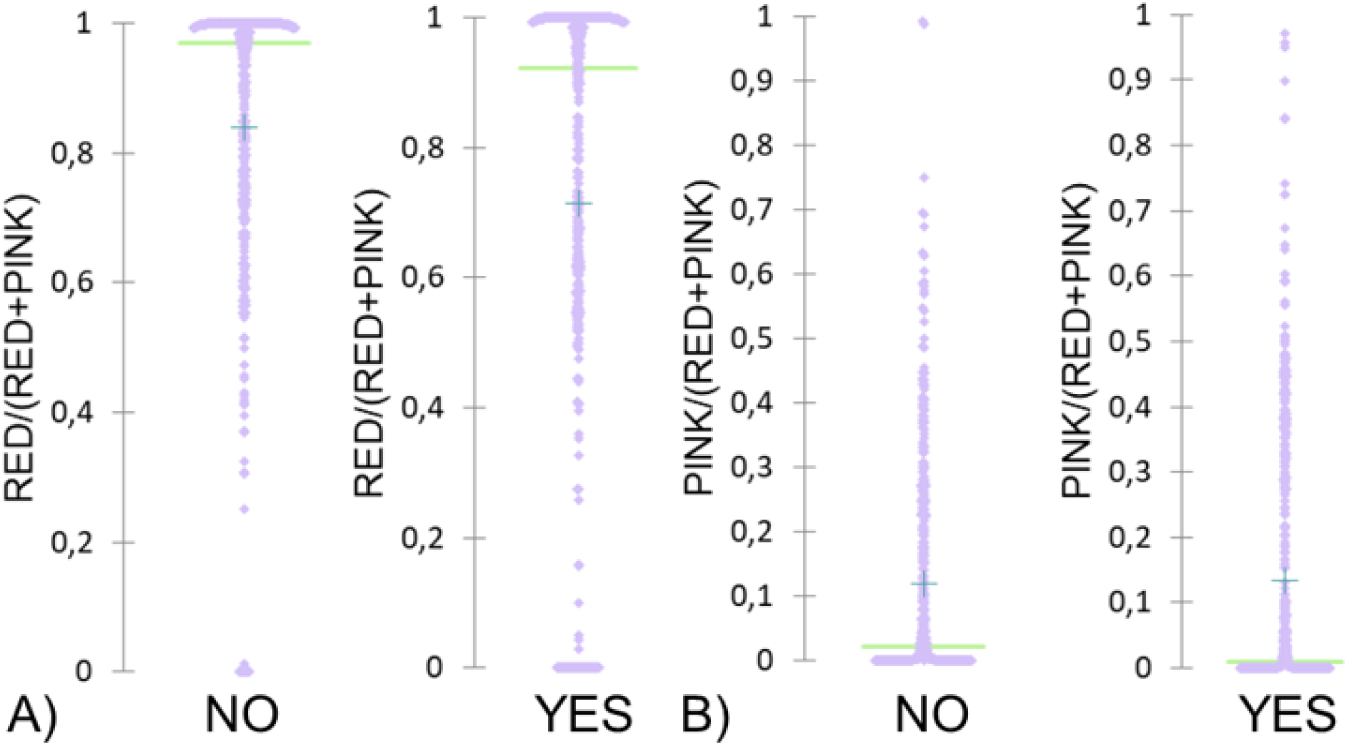
Scattergrams for the classification by TDP-43 pathology from the complete dataset for the variables of (A) cytoplasmic and (B) nuclear proportion.

**Figure B.13:**
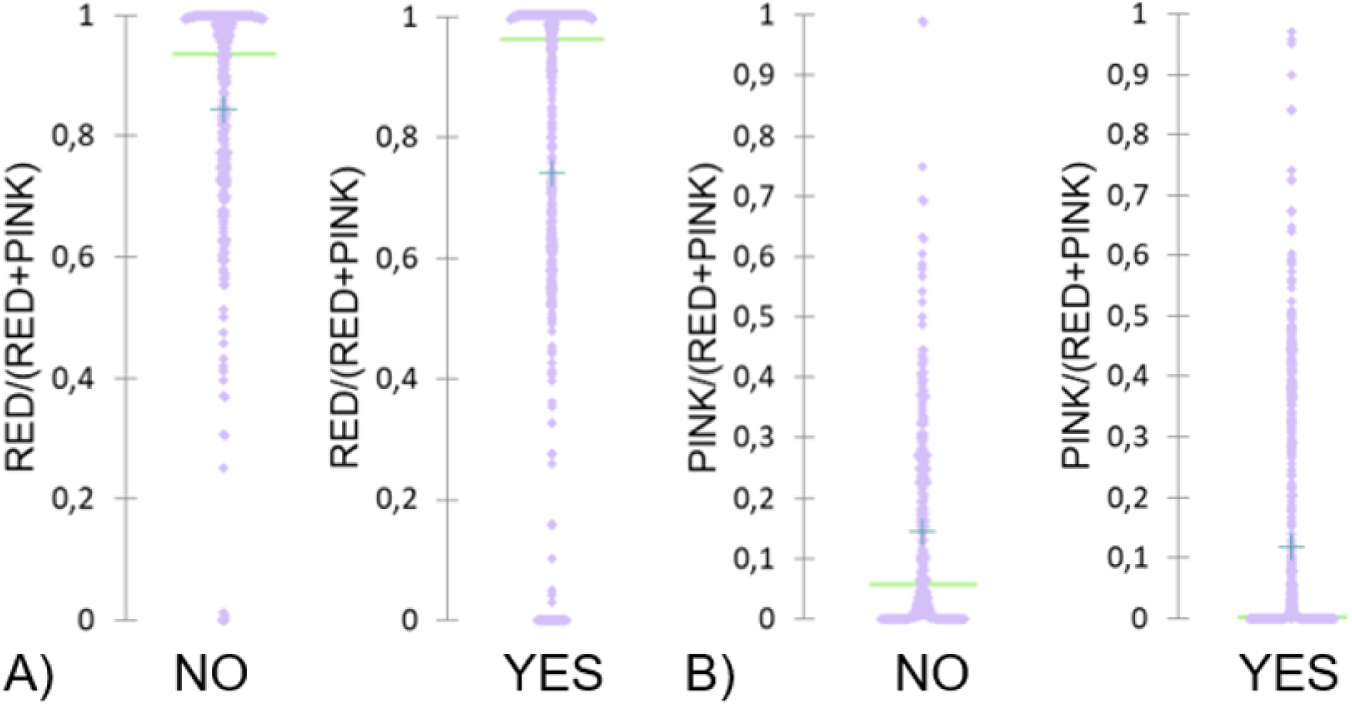
Scattergrams for the classification by ALS from the complete dataset for the variables of (A) cytoplasmic and (B) nuclear proportion.

Figure B.14 displays scattergrams of the classification by grades of TDP-43 pathology from the complete dataset, and in Figure B.15 for the simplified dataset.

**Figure B.14:**
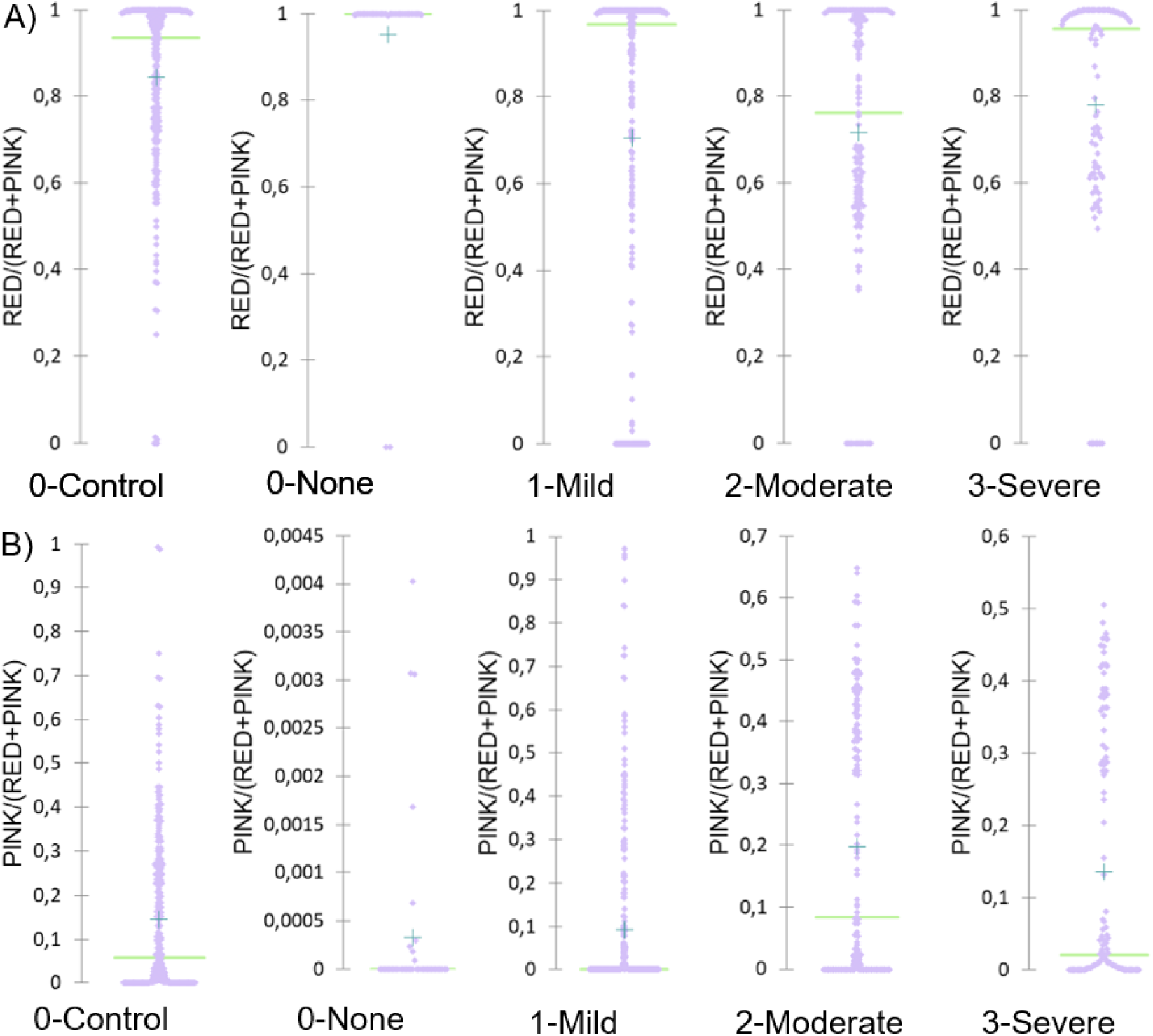
Scattergrams for the classification by grades of TDP-43 pathology from the complete dataset for the variables of (A) cytoplasmic and (B) nuclear proportion.

**Figure B.15:**
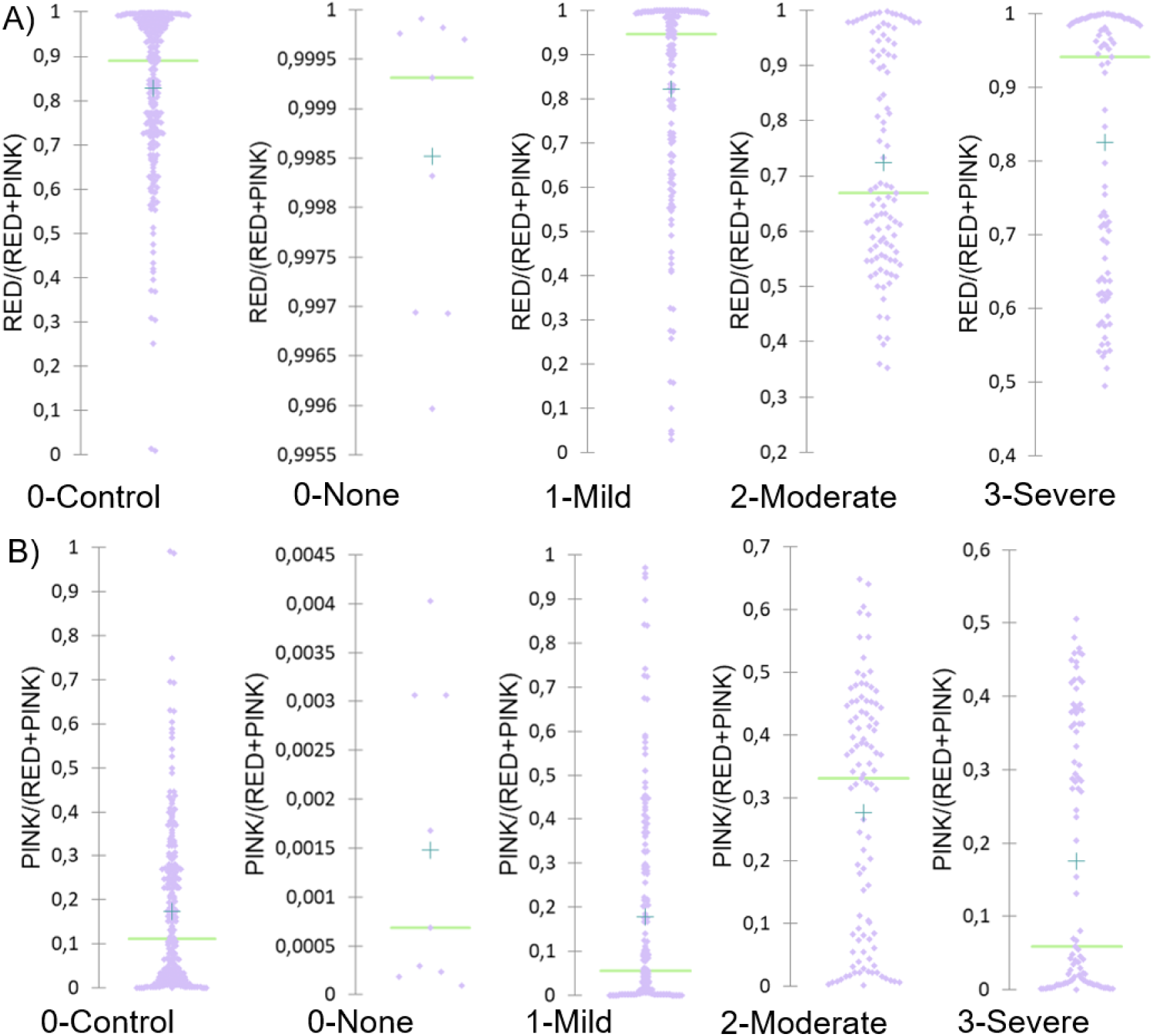
Scattergrams for the classification by grades of TDP-43 pathology from the simplified dataset for the variables of (A) cytoplasmic and (B) nuclear proportion.

Figure B.16 and Figure B.17 illustrate different scattergrams for the classification by TDP-43 pathology and ALS, from the simplified dataset.

**Figure B.16:**
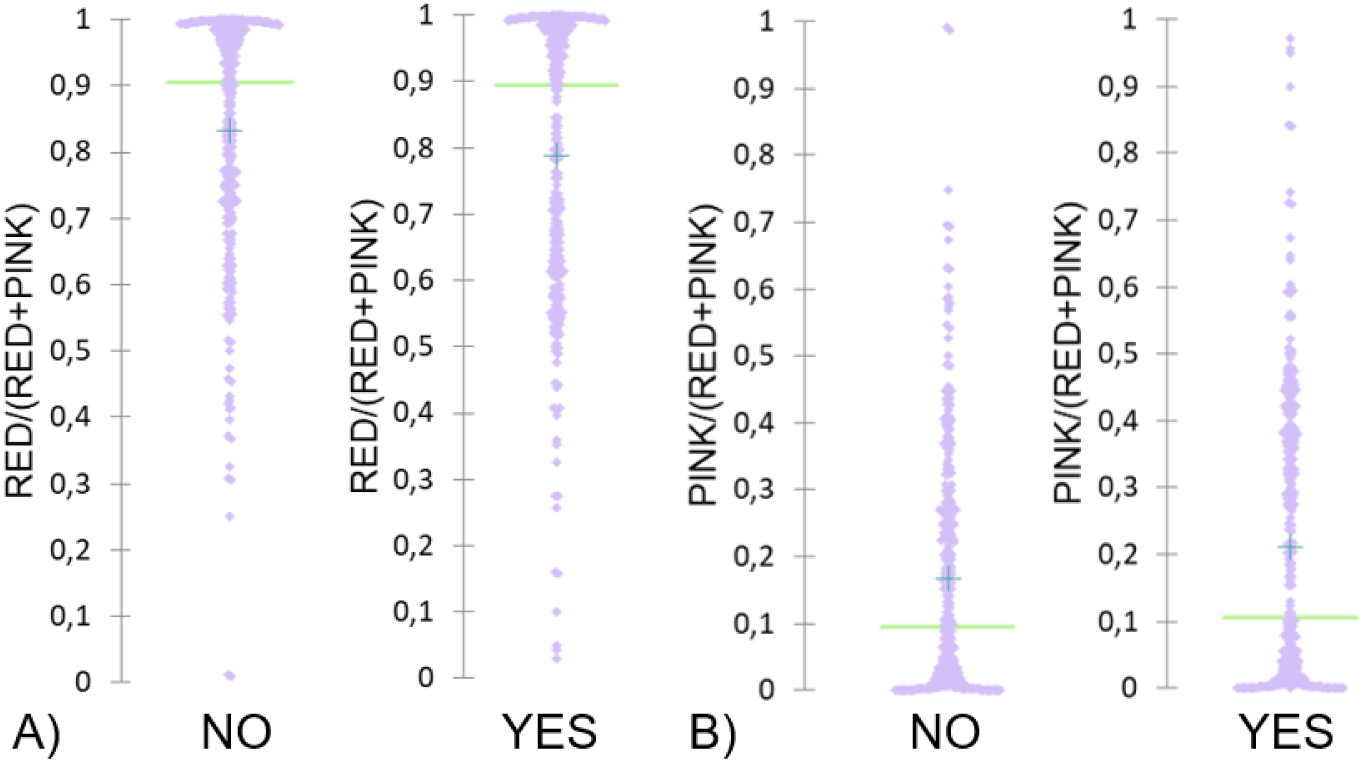
Scattergrams for the classification by TDP-43 pathology from the simplified dataset for the variables of (A) cytoplasmic and (B) nuclear proportion.

**Figure B.17:**
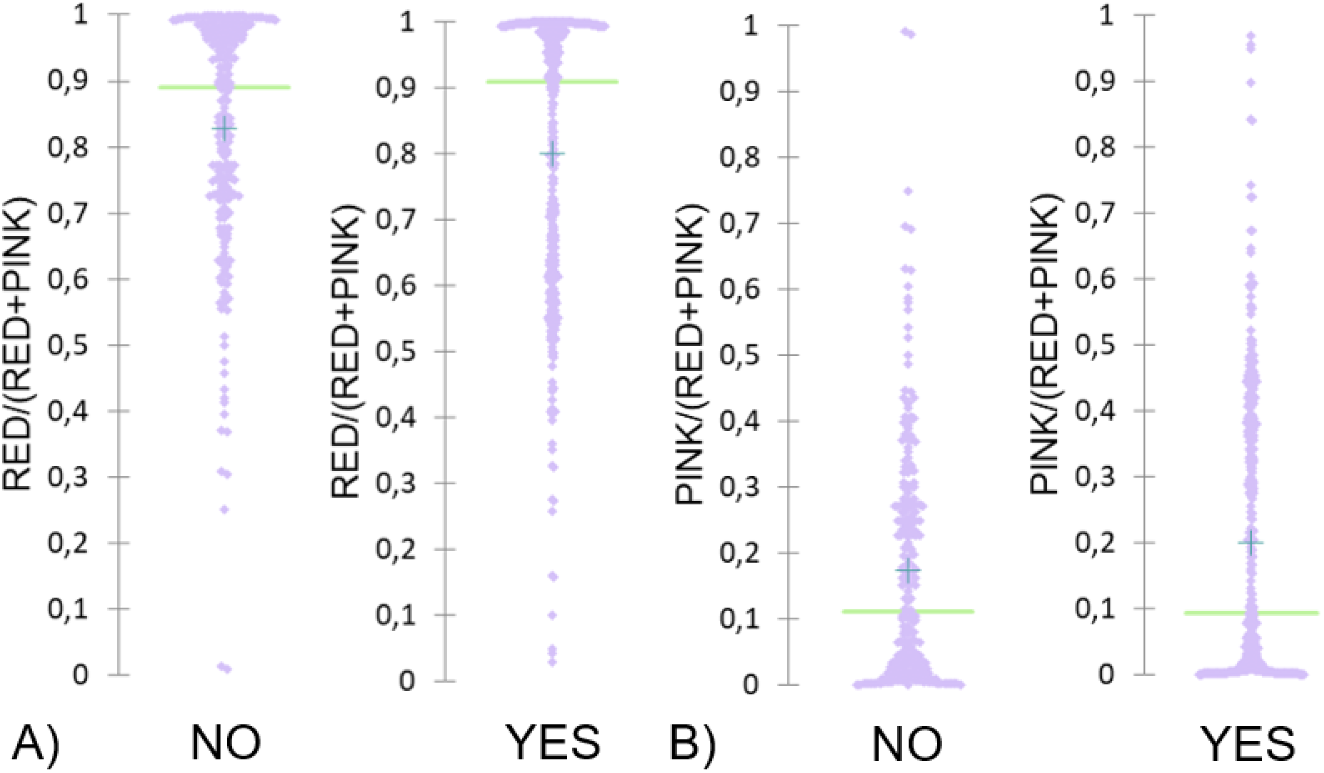
Scattergrams for the classification by ALS from the simplified dataset for the variables of (A) cytoplasmic and (B) nuclear proportion.

